# Reporting patterns of adverse drug withdrawal events using individual case safety reports in United States and European databases

**DOI:** 10.64898/2026.06.15.26355690

**Authors:** Zakir Khan, Ann Sinéad Doherty, Caroline McCarthy, Kieran Dalton, Katharina Tabea Jungo, Emily Reeve, Frank Moriarty

## Abstract

**Introduction:** Adverse drug withdrawal events (ADWEs) are a key safety concern with deprescribing but are infrequently reported in trials. Although pharmacovigilance systems have advanced our understanding of medication-related harms, it is unclear how extensively these systems have been used for ADWEs.

**Objectives:** To examine the reporting patterns of ADWEs for all drugs recorded in United States and European pharmacovigilance databases between 2004 and 2023.

**Methods:** A retrospective study was conducted using two pharmacovigilance databases, the publicly available FDA-FAERS dataset and EMA-EV Level 2A (individual-level) dataset. ADWE cases were identified using relevant MedDRA preferred terms. Data on patient characteristics, reporter type, drugs, indication, ADWE outcomes, dechallenge/rechallenge, seriousness criteria, time to onset, duration, and causality were summarised.

**Results:** A total of 158,505 ADWE reports were analysed (FDA-FAERS: 145,514; EMA-EV: 12,987), with mean ages of 46.1±17.8 (FDA; 55.3% female) and 45.5±21.9 years (EMA; 57.1% female). The frequently reported drug classes were opioids (FDA: oxycodone, 29.8%; EMA: buprenorphine, 19%), antidepressants (FDA: duloxetine, 32%; EMA: venlafaxine, 25.9%) and gabapentinoids (FDA: pregabalin, 6.7%; EMA: pregabalin, 6.0%). The most common adverse outcomes were other serious medical conditions (FDA=63.9%; EMA=46.0%), hospitalisation (FDA=15.9%; EMA=28.3%), and disability (FDA=13.3%; EMA=6.2%) and these outcomes varied significantly based on sex and age group (*p*<0.05).

**Conclusions:** This study provides novel evidence of reporting patterns and characteristics of ADWEs across drugs in pharmacovigilance data. These findings emphasise that adverse drug reaction reporting systems need to accommodate ADWEs (i.e., clarity on terminologies, dechallenge/rechallenge, causality assessment) to effectively capture ADWE-related data to support evidence-based deprescribing practices for better patient safety.

## 1. Introduction

Medication harm is a significant and growing challenge for health systems, designated as the focus of the current World Health Organisation (WHO) Global Patient Safety Challenge [1]. Adverse drug reactions (ADRs) are a subset of adverse drug events (ADEs) and defined as harmful and unintended responses to a drug that happen at a normal dosage used to prevent, diagnose, treat disease, or alter physiological function [2]. Evidence on drug safety from the drug development phase is often incomplete due to the short duration of drug exposure, small samples, and limited population diversity in clinical trials [3, 4]. In routine care, suspected ADRs that occur and are detected are captured via the pharmacovigilance system (post-marketing surveillance program) based on spontaneous reporting of adverse events to generate evidence on safety [4, 5]. The goal of drug post-marketing surveillance programs is to identify and reduce the possibility of medication-related harm [5, 6]. Most countries designate an official pharmacovigilance agency to monitor drug post-marketing surveillance [7]. The Food and Drug Administration (FDA), with its FDA Adverse Event Reporting System (FDA-FAERS) database, supervises the post-marketing safety of drugs in the United States (US) [8]. Similarly, the European Medicines Agency EudraVigilance (EMA-EV) collects ADRs for all approved drugs in the European Economic Area (EEA) [9]. These databases are among the world’s largest collections for passively reported ADRs, including a wide range of patient populations [3, 10, 11].

Adverse drug withdrawal events (ADWEs) are a subset of ADRs (classified as type E reactions – withdrawal/end of use) [12], and refer to clinical symptoms or signs that arise after a drug is discontinued or reduced [13–17]. They can be categorised into physiological withdrawal reactions or the return of medical conditions [10, 15]. Physiological ADWEs are typically short term and more likely to be experienced immediately, whereas the return of medical conditions (also referred to as relapse or recurrence) can be long term and represent re-emergence of the original illness or disease being treated [17–19]. Therefore, ADWEs can manifest as the same symptoms that the medication is used to treat, or as new and unrelated symptoms. ADWEs can range from mild to severe, sometimes requiring the need for hospitalisation [15, 17, 20, 21] and may also be fatal [22, 23]. Any medication has the potential to lead to return of the medical condition and physiological ADWEs have been reported with many medications, including benzodiazepines, antihypertensives, antidepressants, antipsychotics, opioids, antianginals, barbiturates, corticosteroids and proton pump inhibitors [5, 15, 24–27]. However, little is known about the incidence and presentation of ADWEs.

Although the pharmacovigilance system has been used to advance our understanding of medication safety and ADRs caused by prescribing of medications, there appears to be limited use of this system for ADWEs caused by deprescribing (which includes discontinuing or reducing potentially unnecessary or harmful medications) [28]. The limited studies evaluating ADWE reports in pharmacovigilance data have generally focused on a single drug [29–31], or specific drug classes [5, 32, 33], resulting in fragmented evidence and highlighting the need for further investigation to provide a comprehensive assessment of ADWEs. Moreover, there is a lack of evidence on several key aspects of ADWEs, including time to onset, duration, seriousness, causality assessment, and information on typical actions following an ADWE. As ADWE occurrence is a key safety concern during deprescribing, which is infrequently reported in trials [18], strengthening the evidence on ADWEs may help address this barrier and support deprescribing implementation in practice [34]. Therefore, the current study aimed to explore the reporting patterns of ADWEs of all drugs documented in US and European pharmacovigilance databases between 2004 and 2023.

## 2. Methods

### 2.1. Ethical approval and protocol registration

This study was approved by the RCSI University of Medicine and Health Sciences Research Ethics Committee (Reference Number: REC202411011; Dated: 27/01/2025). The study protocol was preregistered on the Open Science Framework at the stage of data cleaning but before any analysis (https://doi.org/10.17605/OSF.IO/UTCWB) [35].

### 2.2. Study design and data source

A retrospective descriptive study of individual case safety reports was conducted in two pharmacovigilance databases (FDA-FAERS and EMA-EV). The FDA-FAERS, which was designed to support the FDA’s post-marketing safety assessment, includes data from ADRs, ADEs, and medication error reports submitted by consumers, healthcare professionals, pharmaceutical manufacturers, and study sponsors. FDA-FAERS data is accessible via its Public Dashboard, which publishes quarterly data extract files (publicly available open data) [36]. The FDA-FAERS and Legacy adverse events reporting system (LAERS) quarterly data extracts were downloaded from the FDA’s website for 2004-2023. The structural and variable-level differences between LAERS and FAERS are presented in **S*upplementary file 1***. Similarly, the EMA-EV database is a repository of information on reports related to ADRs, ADEs, and medication errors submitted by consumers, healthcare professionals, clinical trial sponsors, marketing authorisation holders, and national competent authorities across countries in the European Economic Area. The EMA provides access to ADR data at different levels. Anonymised individual case safety reports were obtained from the EMA-EV following approval of a Level 2A data request. The Level 2A data required for this study consists of detailed patient characteristics, drug information, and reaction descriptions. Details about the descriptions of different variables (FDA-FAERS and EMA-EV) used in this study are provided in ***Supplementary file 2 (Table S2.1 and S2.2)*.**

We conducted analysis separately in both FDA-FAERS and EMA-EV datasets over the same timeframe for comparability. All ADWE report records for any drug from 2004-2023 (20 years) was obtained. The year 2004 was selected as the start of the data inclusion window, corresponding to the beginning of publicly available FDA-FAERS coverage (January 2004) [36].

### 2.3. Identification of ADWEs in databases

ADWEs relating to all drugs (over-the-counter drugs, prescription-only drugs, controlled drugs) were included in the analysis. Events were identified based on the Medical Dictionary for Regulatory Activities (MedDRA) Preferred Terms (PTs, single medical concepts for symptoms, signs, diagnoses, procedures, or other medical characteristics), an internationally standardised terminology used for coding ADRs [37, 38]. Reports are coded by trained individuals with medical backgrounds who are responsible for managing the databases with MedDRA PTs to identify the type of reaction [11, 29]. We identified report records relating to ADWEs as those with a PT containing “withdraw”, and specific irrelevant terms were excluded (i.e. alcohol withdrawal syndrome, drug withdrawal syndrome neonatal, drug withdrawal maintenance therapy etc). Details of the different relevant and non-relevant PTs in the EMA-EV and FDA-FAERS databases are listed in ***Supplementary file 2 (Table S2.3)***.

The FDA-FAERS classifies drugs as “primary suspect” or “secondary suspect” (representing the main drug suspected to be associated with the reported events for the reported PT term) and EMA-EV as “suspect,” without distinguishing between levels of attribution (e.g., primary vs. secondary). We included all drugs reported by the FDA-FAERS as either “primary suspect” or “secondary suspect” in the main analysis, aligning with the EMA-EV’s broader description of “suspect” [39]. This approach helps to minimise potential underestimation of cases where an FDA-classified “secondary suspect” drug may still be causally associated with the reported adverse events. Previous studies have also used a similar approach with FDA-FAERS data [40, 41].

### 2.4. Data deduplication process

The FDA-FAERS and EMA-EV datasets were imported into Stata (version 18), where deduplication was performed. The FDA-FAERS dataset required cleaning to identify and remove potential duplicate reports before analysis. While the EMA applies a procedure for deduplication [42, 43], they note that provided datasets may contain duplicate records. In both databases, the same case can be reported by multiple sources and there is a possibility of multiple entries and duplicate records. Similarly, each individual report may be associated with multiple reporters, ADRs, and medicinal products. To ensure consistency, we retained only the most recent report of a case, ensuring only a single report was retained for each case. After deduplication, one report may contain multiple drugs and/or multiple ADWEs (e.g., as more than one drug may be reported as “suspect”). The data cleaning and deduplication process description is available in ***Supplementary file 2*** (***Table S2.4)*** and relevant scripts used for cleaning are available from ***Zenodo*** [44].

### 2.5. Drug cleaning and mapping

To ensure drug name consistency, we extensively cleaned the FDA-FAERS drug files and mapped brand names to their generic equivalents. We used RxNorm [45] via RxNav (a browser for multiple drug information sources) and Drugs@FDA (the FDA-approved list of products for human use) [46] to map brand names to their corresponding generic active ingredients. The drug cleaning and mapping description is available in ***Supplementary file 2 (Table S2.5)*** and the script is available on ***Zenodo*** [44]. In the EMA-EV, we used the already cleaned drug name variable provided by EMA-EV, which represents the standardised, non-proprietary name of the active product ingredients. Drugs were categorised according to the WHO-Anatomical Therapeutic Chemical (ATC) classification system using 5th-level ATC codes [47]. Moreover, we extracted all drugs from each database and assigned individual drugs to their respective drug classes. Frequencies and percentages were then calculated based on these drug classes.

### 2.6. Statistical analysis

Descriptive analysis (frequency, percentage, means, standard deviation) was used to summarise ADWE report characteristics. We summarised ADWE information from both databases, including patient characteristics (age group and sex), type of reporter, country, year of reporting, ADWE characteristics, seriousness criteria, dechallenge and rechallenge information, time to onset of ADWEs from starting the drug (calculated in the FDA-FAERS= event_dt – drug start date; EMA-EV = start date of reaction – start date of drug) and suspected drug/drug classes details and indication. Moreover, additional details only provided for EMA-EV were summarised: duration of the ADWEs, actions taken with the drug, results of causality assessment (and methods used for this assessment), and outcome of ADWEs at the time of last observation. Although the interpretation of dechallenge and rechallenge is ambiguous for ADWE reports, we summarised this data using the database definitions for these variables and categories. Dechallenge is defined by the FDA-FAERS as “*whether the reaction abated when drug therapy was stopped*” and by the EMA-EV as “*action taken with suspected drug (e.g. drug withdrawn or dose reduced)*” together with “*outcome of reaction/event at the time of last observation*”. Additionally, rechallenge is defined by FDA-FAERS as “whether the reaction recurred when drug therapy was restarted” and by the EMA-EV as “*did the reaction recur on re-administration*”. Further details are available in ***Supplementary file 2*** (***Table S2.6)*.**

Chi-square (χ²) tests were used to assess associations between seriousness criteria and age group (paediatric: 0-17 years, adult: 18-64 years, older adult: ≥65 years) and sex (female, male). Cases with non-specified age and sex groups were excluded from the chi-square analyses to ensure consistency and validity in group comparisons. Statistical significance was considered as *p*<0.05. Logistic regression analyses were also performed to explore the independent relationship with age group and sex and the likelihood of each seriousness outcome among ADWE reports, with results presented as odds ratios (ORs), 95% confidence intervals (CIs), and *p*-values. Stata software (version 18) was used for data management, cleaning, and statistical analysis. Graphs were generated using R (version 4.5.0) with the ggplot2 package.

## 3. Results

### 3.1. ADWE case details in both databases

A total of 158,505 ADWE reports were analysed (FDA-FAERS: 145,514; EMA-EV: 12,987), with mean patient ages of 46.1±17.8 (FDA-FAERS) and 45.5±21.9 years (EMA-EV). In both databases, the proportion of these reports in adults (aged 18-64 years) was higher (FDA-FAERS=30.1% and EMA-EV=21.9%) compared to older adults (FDA-FAERS=5.8% and EMA-EV=4.7%). Where sex was specified, ADWEs were more commonly reported in females versus males (FDA-FAERS=55.3% versus 44.7%; EMA-EV=57.1% versus 42.9%). Considering the type of reporter, only one reporting variable (occupation/qualification as described in both datasets) was reported, this was mainly consumers (41.5%), followed by lawyer (27.7%) and physician (14.0%) in FDA-FAERS. In EMA-EV, consumers or other non-health professionals (40.5%), physician (34.2%) and other healthcare professionals (13.7%) were the leading type of reporters. Pharmacists accounted for the lowest proportion of submitted reports in both databases (FDA-FAERS=2.3%; EMA-EV=11.5%) **(Table 1)**. Additional variable available in FDA-FAERS and EMA-EV about reporting sources/sender types are also provided in ***Supplementary file 2, Table 2.7*.**

**Table 1:** ADWE case details in the FDA-FAERS and EMA-EV databases.

| Variables | FDA-FAERS n (%) | EMA-EV n (%) |
| --- | --- | --- |
| Age (mean ± standard deviation) | 46.1 ± 17.8 years | 45.5 ± 21.9 years |
| **Age group** |  |  |
| Paediatrics | 3114 (2.1) | 269 (2.1) |
| Adult (18-64) | 43833 (30.1) | 2842 (21.9) |
| Older adults (65+) | 8423 (5.8) | 613 (4.7) |
| Not specified | 90144 (61.9) | 9263 (71.3) |
| **Sex** |  |  |
| Female | 74163 (51.0) | 7,153 (55.1) |
| Male | 59942 (41.2) | 5,382 (42.4) |
| Not specified | 11409 (7.8) | 452 (3.5) |
| **Type of reporter \*** |  |  |
| Consumer | 60329 (41.5) | - |
| Lawyer | 40385 (27.7) | - |
| Consumers or other non-health professionals | - | 6,165 (40.5) |
| Physician | 20329 (14.0) | 5,203 (34.2) |
| Other healthcare professional | 14921 (10.2) | 2,094 (13.7) |
| Pharmacist | 3436 (2.3) | 1,751 (11.5) |
| Not specified | 6114 (4.2) | - |
| **Total** | **145,514** | **12,987** |
| **Country of the reporter** |  |  |
| United States | 113,523 (78.0) | - |
| United Kingdom | 8,343 (5.7) | 2,439 (18.8) |
| Canada | 2,883 (2) | - |
| France | 2,520 (1.7) | 3,624 (27.9) |
| Germany | 1,890 (1.3) | 2,674 (20.6) |
| Netherlands | 610 (0.4) | 1,035 (8.0) |
| Sweden | 367 (0.2) | 599 (4.6) |

\* Type of reporter (i.e. consumer or occupation/qualification of the reporter) reported as described in both datasets (see Supplementary File 2-Table S2.1). EMA-EV data allows more than one type of reporter to be recorded.

**Table 2:**
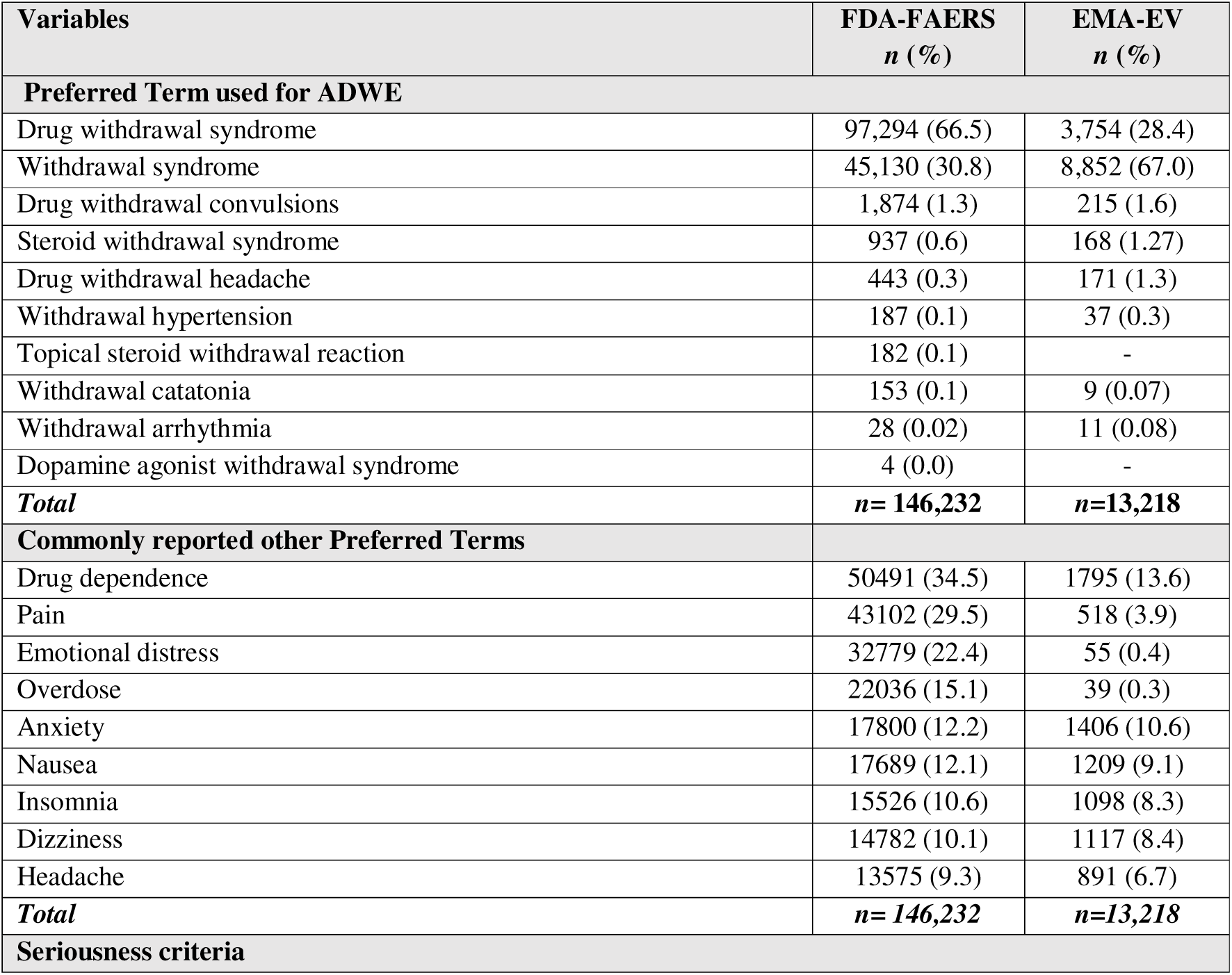
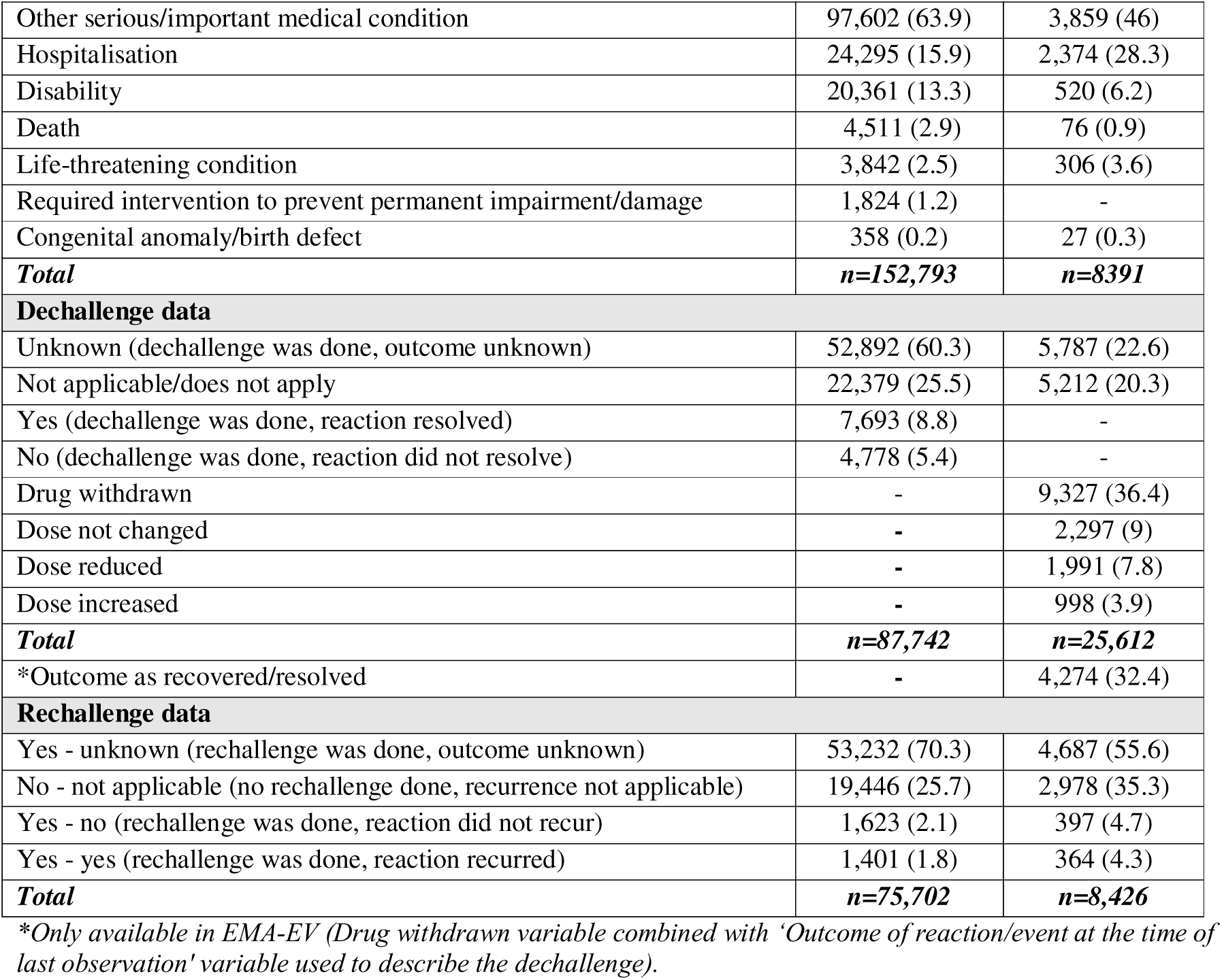
Characteristics of ADWEs in both databases.

In the FDA-FAERS database, the majority of reports were submitted from the United States (78%), with a smaller number of reports included from other countries (e.g. United Kingdom=5.7%, Canada=2.0%). The EMA-EV reports were most frequently from France (27.9%), Germany (20.6%), and the United Kingdom (18.8%) **(Table 1)**. Complete details of country-level distribution of ADWEs reports in FDA-FAERS and EMA-EV databases are listed in ***Supplementary file 3 (Table 3.1 and 3.2)*.**

**Table 3:** Top drug classes and commonly reported drugs in FDA-FAERS and EMA-EV databases.

| Drugs | FDA-FAERS<br>n (%) | Drugs | EMA-EV<br>n (%) |
| --- | --- | --- | --- |
| **Opioids** | **63168 (43.4)** | **Opioids** | **3990 (22.7)** |
| Oxycodone | 18824 (29.8) | Buprenorphine | 759 (19.0) |
| Hydrocodone acetaminophen | 7878 (12.5) | Tramadol | 669 (16.8) |
| Morphine | 7656 (12.1) | Fentanyl | 567 (14.2) |
| **Antidepressants** | **32092 (22)** | **Antidepressants** | **3231 (18.4)** |
| Duloxetine | 10274 (32.0) | Venlafaxine | 838 (25.9) |
| Paroxetine | 9585 (29.9) | Paroxetine | 489 (15.1) |
| Venlafaxine | 5221 (16.3) | Duloxetine | 374 (11.6) |
| **Gabapentinoids** | **9744 (6.7)** | **Gabapentinoids** | **1058 (6)** |
| Pregabalin | 8463 (86.9) | Pregabalin | 927 (87.6) |
| Gabapentin | 1281 (13.1) | Gabapentin | 131 (12.4) |
| ***Hypnotics and sedatives*** | **6436 (4.4)** | ***Hypnotics and sedatives*** | **2078 (11.8)** |
| Alprazolam | 1565 (24.3) | Alprazolam | 320 (15.4) |
| Clonazepam | 1490 (23.2) | Lorazepam | 267 (12.8) |
| Lorazepam | 843 (13.1) | Oxazepam | 256 (12.3) |
| ***Muscle relaxants*** | **4512 (3.1)** | ***Muscle relaxants*** | **284 (1.6)** |
| Baclofen | 4305 (95.4) | Baclofen | 273 (96.1) |
| Carisoprodol | 98 (2.2) | Tizanidine | 6 (2.1) |
| Tizanidine | 87 (1.9) | Botulinum toxin | 2 (0.7) |
| ***Antipsychotics*** | **4471 (3.0)** | ***Antipsychotics*** | **905 (5.1)** |
| Quetiapine | 1519 (34.0) | Quetiapine | 251 (27.7) |
| Olanzapine | 562 (12.6) | Olanzapine | 135 (14.9) |
| Aripiprazole | 496 (11.1) | Clozapine | 117 (12.9) |
| ***Drugs used in addictive disorders*** | **2335 (1.6)** | ***Drugs used in addictive disorders*** | **362 (2)** |
| Varenicline | 893 (38.2) | Varenicline | 159 (43.9) |
| Naltrexone | 877 (37.6) | Nicotine | 63 (17.4) |
| Nicotine | 530 (22.7) | Nalmefene | 59 (16.3) |
| ***Antiepileptics*** | **2052 (1.4)** | ***Antiepileptics*** | **367 (2.1)** |
| Vigabatrin | 528 (25.7) | Valproic acid | 92 (25.1) |
| Lamotrigine | 386 (18.8) | Lamotrigine | 69 (18.8) |
| Valproic acid | 296 (14.4) | Carbamazepine | 50 (13.6) |
| ***Antihistamines*** | **1445 (1)** | ***Antihistamines*** | **191 (1.1)** |
| Cetirizine | 1111 (76.9) | Cetirizine | 94 (49.2) |
| Diphenhydramine | 121 (8.4) | Levocetirizine | 34 (17.8) |
| Fexofenadine | 56 (3.9) | Promethazine | 13 (6.8) |
| ***Psychostimulants*** | **1389 (0.9)** | ***Psychostimulants*** | **192 (1.1)** |
| Methylphenidate | 469 (33.8) | Methylphenidate | 91 (47.4) |
| Lisdexamfetamine | 279 (20.1) | Methylenedioxymethamphetamine | 30 (15.6) |
| Amphetamine dextroamphetamine | 269 (19.4) | Lisdexamfetamine | 18 (9.4) |
| ***Corticosteroids n (%)*** | **1167 (0.8)** | ***Corticosteroids*** | **317 (1.8)** |
| Prednisone | 250 (21.4) | Prednisolone | 72 (22.7) |
| Prednisolone | 183 (15.7) | Betamethasone | 62 (19.6) |
| Hydrocortisone | 177 (15.2) | Hydrocortisone | 52 (16.4) |
| ***Antihypertensives n (%)*** | **992 (0.7)** | ***Antihypertensives*** | **193 (1.1)** |
| Clonidine | 323 (32.6) | Clonidine | 31 (16.1) |
| Metoprolol | 79 (8.0) | Metoprolol | 21 (10.9) |
| Propranolol | 79 (8.0) | Guanfacine | 17 (8.8) |

### 3.2. ADWEs reporting trends over time

Analysis of reporting trends from 2004 to 2023 shows that FDA-FAERS had a significant increase in ADWE reports over time, rising from 4,490 reports in 2004 to a peak of 24,316 reports in 2021, followed by a slight decline in 2022–2023. In contrast, EMA-EV demonstrated a more gradual upward trend, with reports increasing from 106 in 2004 to a peak of 1,323 in 2020, and then slightly decreasing in subsequent years **(Figure 1).** More details on ADWE reporting trends over the 20-year period in FDA-FAERS and EMA-EV are presented in ***Supplementary file 3 (Table S3.3)***.

**Figure 1:**
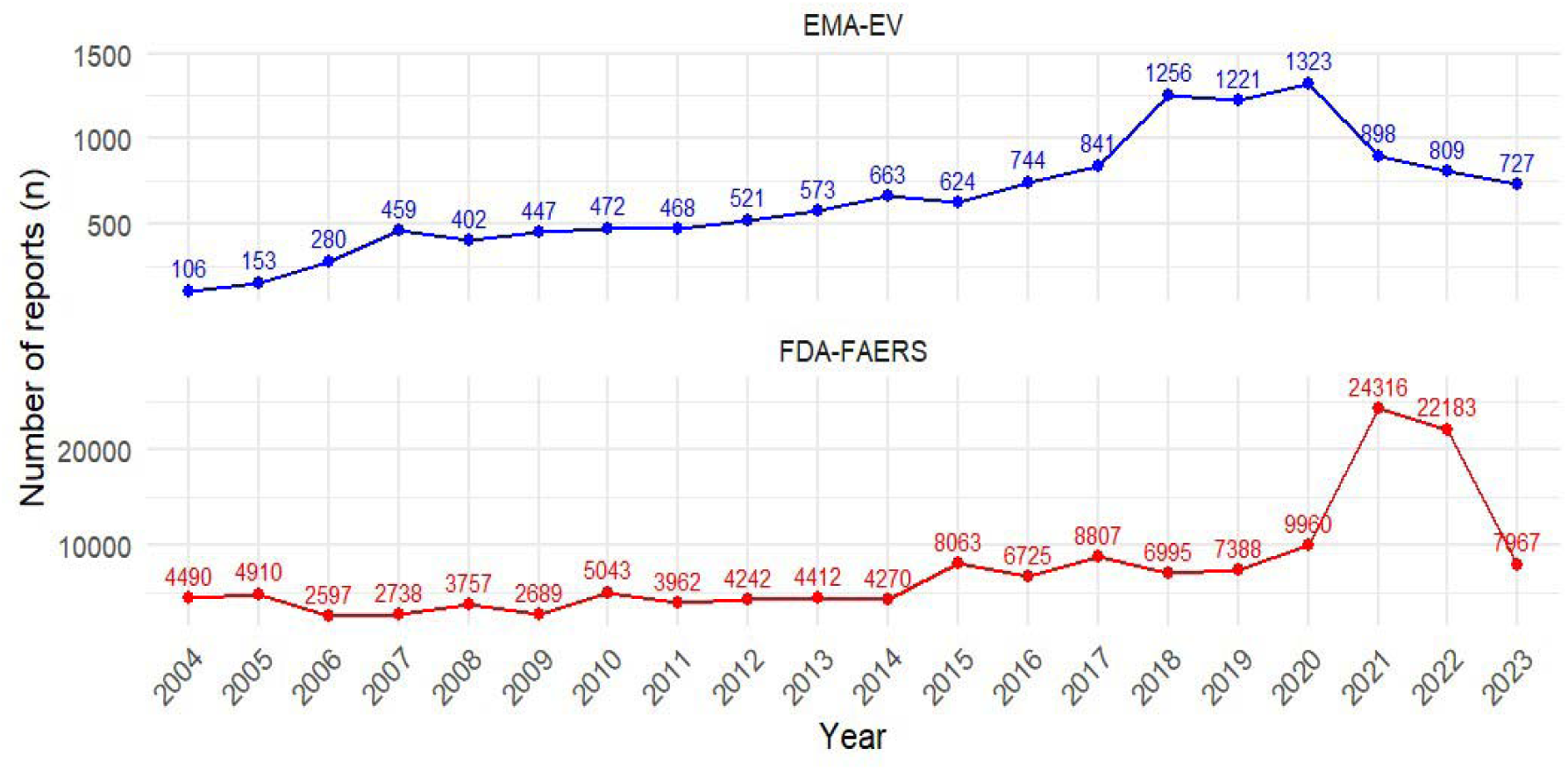
ADWEs reporting trends over time in FDA-FAERS and EMA-EV (2004–2023)

### 3.3. Preferred Terms (PTs) of ADWEs and other PTs

‘Drug withdrawal syndrome’ was the most frequently reported PT in included FDA-FAERS reports (*n*=97,294; 66.5%), whereas ‘withdrawal syndrome’ was the most common in EMA-EV reports (*n*=8,852; 67%). Other PTs, including ‘drug withdrawal convulsions’, ‘steroid withdrawal syndrome’, and ‘drug withdrawal headache’ were reported in both databases (FDA-FAERS <3% combined; EMA-EV <5% combined). Rare PTs, such as ‘withdrawal hypertension’, ‘withdrawal catatonia’, and ‘withdrawal arrhythmia’ were also reported, with some PTs (topical steroid withdrawal reaction and dopamine agonist withdrawal syndrome) reported only in the FDA-FAERS database **(Table 2)**. The most common other PTs included in reports containing ADWE PTs included drug dependence, pain, emotional distress, overdose, and anxiety (see **Table 2**, with full details of the top other PTs available in ***Excel supplementary file E1*** and ***Excel supplementary file E2***).

### 3.4. Seriousness criteria and details on dechallenge and rechallenge

ADWE seriousness was reported in both databases. Hospitalisation due to ADWEs was common, reported in 15.9% (*n*=24,295) of FDA-FAERS cases and 28% (*n*=2,374) of EMA-EV cases, followed by disability (FDA-FAERS=13.3%; EMA=6.2%). However, “other serious/important medical condition” was the most frequently reported category in both databases (63.9% FDA-FAERS vs. 46% EMA-EV) **(Table 2)**. Dechallenge was documented in 74.5% (65,363/87,742) of FDA-FAERS cases (yes = dechallenge was done, reaction resolved; no = dechallenge was done, reaction did not resolve and unknown = dechallenge was done, outcome unknown and 66.8% (17,105/25,612) of EMA-EV cases (unknown/drug withdrawn/dose reduced). Additionally, rechallenge was documented in 74.3% (FDA-FAERS=56,256/75,702) and 64.6% (EMA-EV=5,448/8,426) of cases (**Table 2).**

### 3.5. Onset of ADWEs

Considering onset of ADWEs, only cases with documented date of onset of reaction were included in the analysis (FDA-FAERS: *n*=10,996; 7.5%, EMA-EV: *n*=3,701; 28.5%). There was a bimodal distribution, with ADWEs most commonly occurring either within ≤1 day (FDA-FAERS=28.6%; EMA-EV=15.6%), or >1 year (FDA-FAERS=23.2%; EMA-EV=34.9%) based on reported time to onset, followed by 1-3 months (FDA-FAERS=11.3%; EMA-EV=10.9%) and 6–12 months (FDA-FAERS=9.8%; EMA-EV=13.8%) **(Figure 2 *and supplementary file 3: Table S3.4)***. Moreover, individual results per ADWE PTs are also presented in ***Supplementary file 3 (Tables S3.5 and S3.6)***.

**Figure 2:**
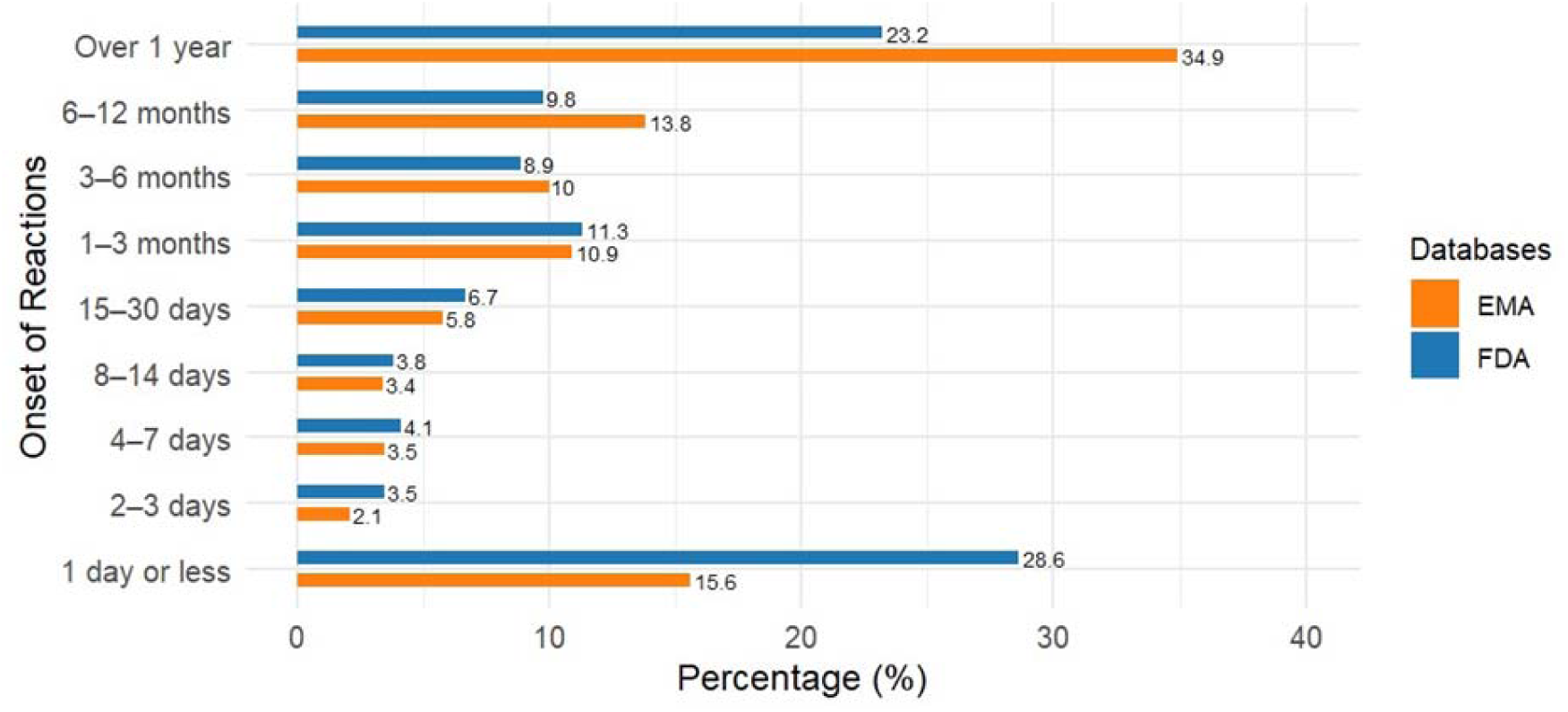
Onset of ADWEs in FDA-FAERS and EMA-EV databases.

### 3.6. Additional ADWE characteristics in EMA-EV

Additional characteristics, including outcome (event at the time of last observation) of ADWEs, duration of reaction, and causality assessment are only available in the EMA database. Outcomes of ADWEs at last observation were unknown in 38.8% (*n*=5,109), recovered/resolved in 32.4% (*n*=4,274), and not recovered in 17.2% (*n*=2,266). A small proportion had recovered with sequelae (*n*=109; 0.8%) or were fatal (*n*=41; 0.3%). Causality assessment was most frequently based on clinical judgment or global introspection (*n*=8,785; 70%), WHO-UMC method (*n*=892; 7.1%) followed by French imputability (*n*=851; 6.8%) and Naranjo (*n*=459; 3.6%). Details about causality assessment approaches are listed in ***Supplementary file 4 (Table S4.1)***. ADWEs were most commonly classified as possible (*n*=5,393; 53.9%) and probable/likely (*n*=2,859; 28.6%), excluding the unassessable/unclassifiable and unknown cases. A relatively small number of reactions were classified as certain/definite (*n*=134; 1.3%) and doubtful (*n*=131; 1.3%). Finally, in EMA-EV, cases are classified using the EU assessment method (with two outcomes: ‘reasonable possibility’ and ‘no reasonable possibility’). This was reported in 1,615 cases, with 80.1% (1,294) classified as reasonably possible (suggesting a causal relationship between the drug and adverse event) **(*Supplementary file 4: Table S4.1)***.

Among 1,403 ADWEs cases with valid duration of reaction data (1,403/12,987, 10.8%), the majority (*n*=893; 63.6%) had a reaction duration of one week or less. The most commonly observed durations of reaction were 2–3 days (*n*=322; 23%), ≤1 day (*n*=303; 21.6%), 4–7 days (*n*=268; 19.1%), and 8–14 days (*n*=213; 15.2%). Moreover, 10.8% (*n*=151) of all ADWEs continued for more than one month, including those that lasted 1–3 months (5.9%) and over one year (1.4%) **(Figure 3 and Supplementary file 4: Table S4.2)**. Drug withdrawal convulsions were mostly short in duration (65.7% ≤1 day). Similarly, withdrawal syndrome and drug withdrawal syndrome cases mostly lasted <1 week (63.8% and 61.8% respectively). Prolonged durations were commonly observed in steroid withdrawal syndrome (44.4% of cases with 1 month to 1 year and 22.2% lasted over a year) **(Supplementary file 4: Table S4.3).**

**Figure 3:**
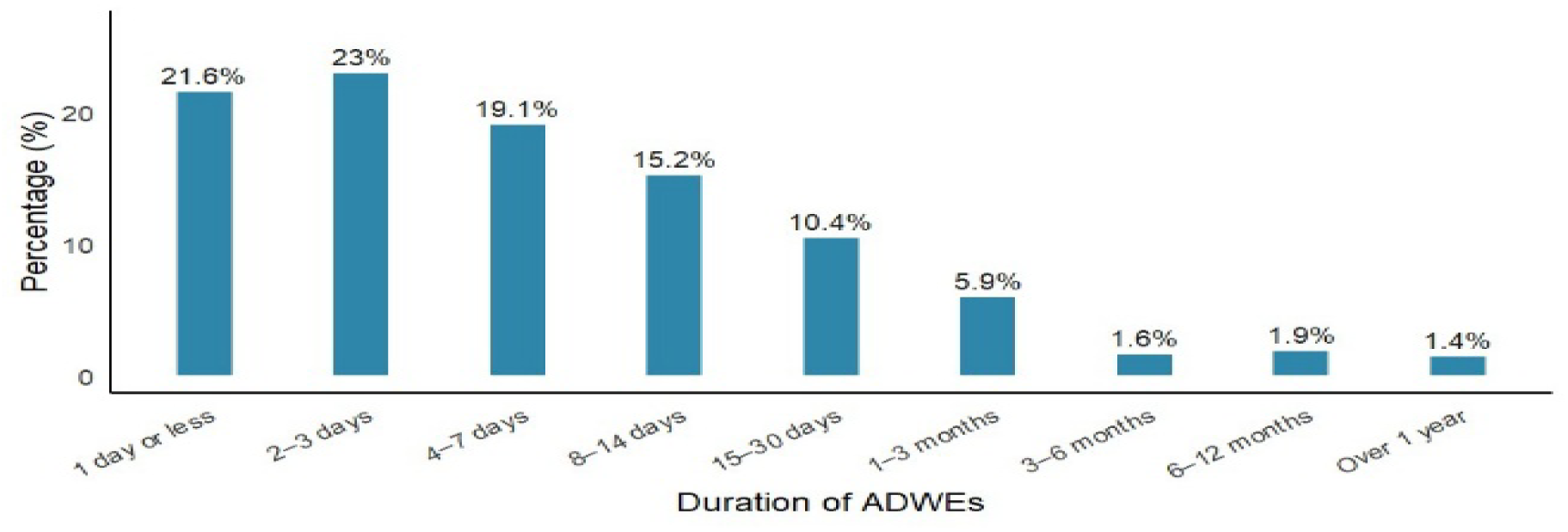
Duration of ADWEs in EMA-EV database.

### 3.7. Top drug classes associated with ADWEs in both databases

A total of 145,552 (1329 unique drugs) drugs from the FDA-FAERS database and 17,525 (1015 unique drugs) from the EMA-EV database were reported in ADWE reports. Among the top drug classes, opioids (FDA-FAERS: 63168/145552=43.4%; EMA-EV: 3990/17525=22.7%) were most frequently reported in both databases followed by antidepressants (FDA-FAERS=22%; EMA-EV=18.4%), gabapentinoids (FDA-FAERS=6.7%; EMA-EV=6%), hypnotics and sedatives (FDA-FAERS=4.4%; EMA-EV=11.8%), muscle relaxants (FDA-FAERS=3.1%; EMA-EV=1.6%), and antipsychotics (FDA-FAERS=3%; EMA-EV=5.1%). Additionally, ADWEs with other drug classes including antiepileptics, drugs used in addictive disorders, antihistamines, psychostimulants, corticosteroids, and antihypertensives were also reported **(Table 3).** More details about different drug classes and common drugs are available in ***Supplementary file 5 (Table S5.1)*.**

### 3.8. Association of seriousness criteria of ADWEs reports with age and sex group

Chi-square analyses of ADWE seriousness criteria revealed significant differences by age and sex in both the FDA-FAERS and EMA-EV databases, detailed in ***Supplementary file 6 (Table S6.1 and S6.2)***. Multivariable logistic regression analyses adjusting for age group and sex were conducted using FDA-FAERS (*n*=54,698;37.6%) and EMA-EV (*n*=2,312;17.8%) reports with complete information on both covariates. Across both datasets, paediatric patients (FDA-FAERS: *OR*=1.52; EMA-EV: *OR*=3.24) and older adults (FDA-FAERS: *OR*=1.29; EMA-EV: *OR*=1.46) had significantly higher odds of prolonged hospitalisation compared with adults. The “Other serious/important medical condition” category also showed significantly higher odds among paediatric patients (FDA-FAERS=1.60; EMA-EV=1.36) and older adults (EMA-FAERS=1.71) relative to adults. Older adults also had significantly higher odds of death (FDA-FAERS=1.35; EMA-EV=1.43). However, both paediatric (FDA-FAERS=0.67, EMA-EV=0.16) and older adults (FDA-FAERS=0.47, EMA-EV=0.50) had significantly lower odds of disabling events relative to adults. Moreover, sex-based analysis showed that male patients compared to females had significantly higher odds of death (FDA-FAERS=2.90; EMA-EV=3.89) followed by hospitalisation (FDA-FAERS=1.41; EMA-EV=1.68) and life-threatening condition (FDA-FAERS=1.19; EMA-EV=2.06) in both databases. No significant sex differences were observed for disability or congenital anomalies in the FDA-FAERS database. However, in the EMA-EV database, male patients had significantly higher odds of congenital anomalies (*OR*=1.63) compared with females **(Table 4).**

**Table 4:**
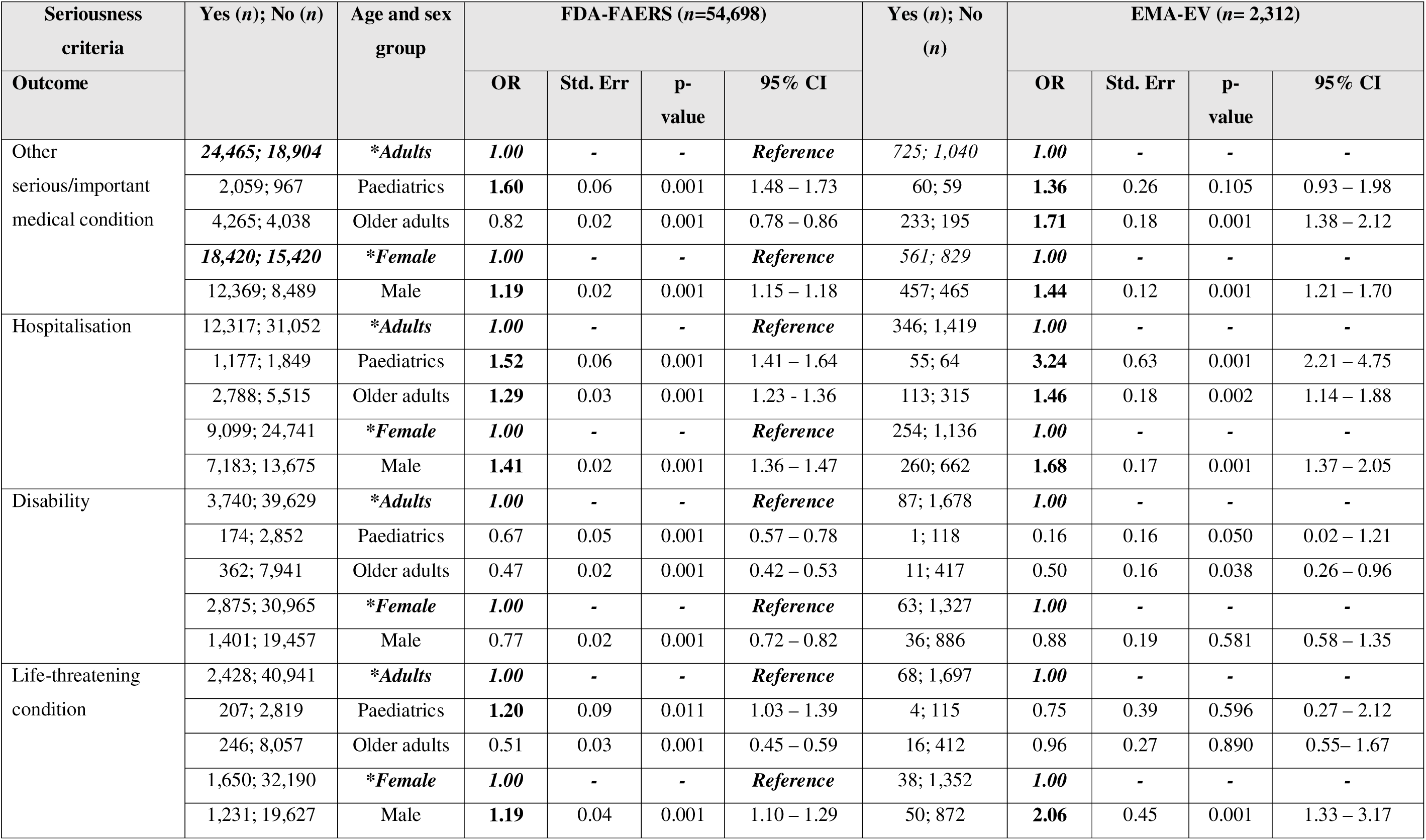
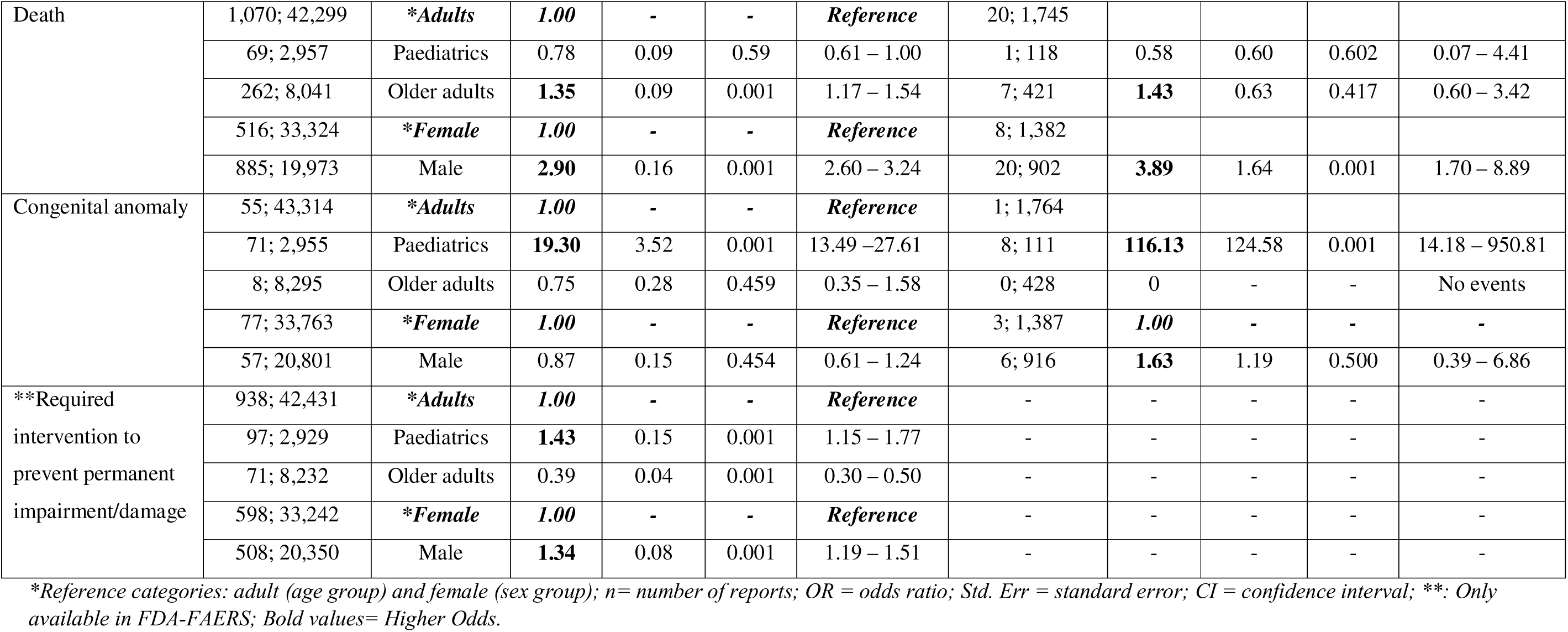
Multivariable logistic regression analysis by age and sex group in FDA-FAERS and EMA-EV databases.

## 4. Discussion

### 4.1. Main Findings

To the best of our knowledge, this study represents the most comprehensive analysis of ADWEs across different drug classes, using data from two international pharmacovigilance databases. No previous study has evaluated ADWEs in relation to patient characteristics (such as age and sex), severity of ADWEs, and other ADR report characteristics.

Our study observed a higher proportion of ADWE reports in females compared to males in both the FDA-FAERS (55.3%) and EMA-EV (57.1%) databases. This female predominance has also been consistently reported in previous pharmacovigilance studies of withdrawal events to specific drug classes, including selective serotonin reuptake inhibitors (SSRIs) [11], antidepressants [5], and antipsychotics [32] across multiple databases (FDA-FAERS, EMA-EV, and WHO/VigiBase). However, differences in reported proportions of females between studies are likely due to methodological variations, as previous studies often focused on selected drug classes [5, 11, 32] or limited relevant PTs (e.g., “drug withdrawal syndrome”) [10]. These findings suggest that females may experience and have ADWEs reported more frequently than males, possibly due to both sex-related biological changes (e.g., differences in pharmacokinetics), and gender-related factors (e.g., healthcare-seeking behaviours and the types of medications prescribed to women versus men) [48, 49]. In contrast, male patients had significantly higher odds of having more serious outcomes compared to females. One possible explanation is underreporting of milder events among males. These findings were also supported by previous studies of ADRs that highlighted variations in seriousness criteria in sex-stratified analysis [48, 50, 51]. The seriousness of the reaction provides important information about the level of risk involved, as serious ADWEs have been associated with longer hospital stays and higher financial costs [52, 53]. Therefore, variations in the reporting frequency and seriousness of ADWEs across sex groups highlight the need to consider these factors during drug discontinuation and suggest that greater emphasis may be needed to encourage the reporting of ADWEs among males. Additional research is needed to clarify the influence of sex-related factors on the occurrence, reporting, and seriousness of ADWEs.

The majority of reports in both databases were submitted by consumers or non-health professionals, followed by healthcare professionals (HCPs), including physicians, pharmacists, and other healthcare providers. Underreporting is a well-recognised limitation of spontaneous reporting systems and has been highlighted in previous studies [54–57]. ADWEs are also likely underreported in pharmacovigilance databases. In this study, HCPs showed lower reporting of ADWEs compared with consumer/non-health professionals. Reporting of ADWEs, similar to ADRs, is an important professional responsibility of all HCPs within the pharmacovigilance system. Therefore, targeted pharmacovigilance training or continuing professional development as highlighted by previous studies on spontaneous ADR reporting and medication error prevention may help enhance rates of HCPs ADWEs reporting [6, 58, 59]. Among HCPs, pharmacists contributed the smallest proportion of submitted reports (FDA-FAERS: 2.3%; EMA-EV: 11.5% overall). Previous studies on ADR reporting also highlighted limited pharmacist involvement in pharmacovigilance activities [6, 10, 60–62]. ADWEs are often more likely to be clinically recognised and diagnosed by physicians, while pharmacists may contribute through the early identification of potential ADWEs. Pharmacists alongside physicians and other healthcare providers play a crucial role in medication safety program, including pharmacovigilance and deprescribing activities [6, 62–64]. Therefore, strengthening multidisciplinary engagement and awareness among all HCPs, including pharmacists, may improve the recognition and reporting of ADWEs in clinical practice.

In this study, a large difference in the number of ADWE reports between the FDA-FAERS and EMA-EV databases is likely to reflect a combination of structural, regulatory, and contextual factors that influence reporting behaviour. These differences may be due to the earlier establishment and longer development of pharmacovigilance infrastructure in the United States, including the introduction of spontaneous reporting systems and the MedWatch programme in the early 1990s, requirements for reporting serious and non-serious cases from all sources (domestic and foreign) in 2001, and the implementation of FDA-FAERS with mandatory electronic reporting and expanded post-marketing safety data collection from 2012 onwards [36, 65, 66]. In contrast, the EMA-EV system was introduced in 2001, with major pharmacovigilance legislation coming into force in 2012 and the introduction of an improved version of the EudraVigilance system in 2017, reflecting a more recent and progressively evolving reporting framework [67–69]. Additionally, differences in reporting volume may also be influenced by regional reporting practices, healthcare system organisation and reporting culture context; for example, increased awareness of opioid-related harms associated with previous opioid epidemic waves in the United States may also be associated with higher reporting [70, 71].

This study also showed an increasing trend in ADWE reporting, with peaks in 2020-2021 and then reports in both databases declined during more recent years. A similar trend was also reported in a previously conducted study in the FDA-FAERS database related to ADWE reports [10]. The increasing trend may be associated with more reporting in general during the COVID-19 pandemic, increased public awareness, enhanced reporting of adverse events, and greater engagement of consumers and healthcare professionals in pharmacovigilance activities [69, 72, 73]. The decline in ADWE reports observed after 2021 may reflect several factors, including the completion of mass vaccination campaigns, normalisation of routine pharmacovigilance activities following intensified monitoring during the COVID-19 pandemic, and decreased public and professional attention to reporting. However, there was an increase in reporting in the EMA-EV in 2018 (pre-COVID-19) compared with FDA-FAERS. This rise may be attributed to the introduction of an improved version of the EudraVigilance system in 2017 in Europe, which simplified the submission of suspected adverse reactions and also a requirement to report non-serious cases, whereas previously only serious cases were included. In 2018, EMA-EV received over 2 million reports of suspected adverse effects, a 37% increase compared with 2017 [68].

In our study, only 7.5% (*n*=10,996) of FDA-FAERS reports and 28.5% (*n*=3,701) of EMA-EV reports included information on time to reaction onset, highlighting substantial limitations in temporal data completeness within spontaneous reporting systems. It is not possible to definitively distinguish between physiological withdrawal reactions and recurrence of the underlying condition, as this detail is not explicitly captured in either database. However, the distribution of reported time-to-event may provide indirect contextual information. Specifically, early-onset events occurring shortly after treatment discontinuation are more consistent with physiological withdrawal mechanisms, whereas markedly delayed onset reports (e.g. >1 year) may be more compatible with recurrence of the underlying condition.

In the present study, we included all relevant PTs containing “withdraw” as per MedDRA in both databases to capture all relevant ADWE reports. This is a strength of the current study compared to previous research that only included the PTs “drug withdrawal syndrome” and “drug withdrawal syndrome neonatal” in the FDA-FAERS database [10], while a study using the EMA-EV database included only four PTs to identify ADWEs [29]. Moreover, two previous studies that analysed the WHO/VigiBase and French Pharmacovigilance databases identified ADWEs using only the PT “withdrawal syndrome” [5, 74]. MedDRA includes a limited set of terms related to drug withdrawal, which may contribute to under-recognition and misclassification of ADWEs. Currently, a “withdrawal” PT indicates only that the reaction occurred after drug discontinuation, without providing information on the specific signs/symptoms, or nature of the reaction (i.e., whether it is a physiological withdrawal reaction or a return of the condition). This differs from typical ADR reporting, where PTs describe the specific signs and symptoms (e.g., insomnia, hypertension, dizziness, anxiety), and suggests a need to more precisely characterise ADWEs within reports.

Dechallenge and rechallenge of drugs are important steps to identify a potential causal relationship between a suspected drug and an adverse reaction [75]. Current ADR reporting forms include “dechallenge” (whether a reaction abated after drug discontinuation or dose reduction) and “rechallenge” (whether a reaction reappeared after reintroduction) variables using yes/no/unknown and does not apply options. However, these fields used to support causality assessment for ADRs during prescribing may be ambiguous when applied to ADWEs (i.e., reactions that generally occur after drug discontinuation). It is unclear how these variables were interpreted or applied in the context of ADWEs in both databases, and further adaptation or clarification on the meaning of these fields in the context of ADWE reporting would be beneficial [76].

In this study, clinical judgment (global introspection) was the most commonly reported method for causality assessment, followed by the WHO-UMC system, the French imputability method, and the Naranjo algorithm – all methods typically used for assessing ADR causality. A recent systematic review identified six tools/criteria used to report ADWEs in deprescribing trials including the Naranjo probability scale, clinical monitoring, identification through the International Classification of Diseases (ICD) system, classification of ADWEs as a subset of confirmed ADEs, patient/caregiver self-reports, and clinical judgment [18]. However, no single method is universally accepted as the reference standard for causality assessment, as each has its own strengths and limitations [18, 77, 78]. Therefore, there is a need to develop robust ADWE detection criteria that incorporate a causality assessment component to ensure accurate ADWE identification in pharmacovigilance data.

In this study, opioids were the most frequently reported drug class, aligning with findings in a previous pharmacovigilance study of drug withdrawal syndrome and neonatal drug withdrawal syndrome PTs [10]. Opioid ADWEs are a key driver behind continued opioid use, and a barrier to opioid discontinuation [79]. The severity and type of opioid ADWEs varies among different opioid drugs and between patients [80]. Therefore, pharmacovigilance data may help address this by systematically collecting ADWEs across drugs and patient populations, helping clinicians identify which opioids pose higher risks for specific patients and guiding more informed prescribing decisions. ADWEs with psychotropic drugs, including antidepressants (e.g. serotonin-norepinephrine reuptake inhibitors, selective serotonin reuptake inhibitors), benzodiazepines, and antipsychotics were also commonly reported, reflecting their known association with withdrawal phenomena and dependence potential [15, 17, 18, 81–83]. In our study, antidepressants were the second most common drug class among ADWE reports, consistent with previous pharmacovigilance studies focused on antidepressant-related ADWE reports from the WHO database [5, 25], as well as studies on SSRIs using the FDA-FAERS and EMA-EV databases [11] and the French Pharmacovigilance database [74]. Moreover, ADWE cases were also reported for other drug classes, including gabapentinoids, muscle relaxants, antihistamines, corticosteroids, antihypertensives, and proton pump inhibitors. These findings suggest that clinically relevant ADWEs are not limited to centrally acting drugs and may also occur following discontinuation of other drug classes. This highlights the importance of careful monitoring and gradual dose reduction during drug discontinuation [15].

### 4.2. Strengths and Limitations

Pharmacovigilance data are an important and reliable source to monitor drug safety, particularly in an area where large-scale epidemiological data are limited and clinical decisions rely mainly on expert opinion or small studies. A key strength of this study is the analysis of two large, independent regulatory databases (FDA-FAERS and EMA-EV), which enhances the robustness and external validity of the findings. The use of both datasets enables cross-system comparison of ADWE patterns across different regulatory frameworks and geographical regions, thereby improving the generalisability of the results. This study also provides detailed step-by-step documentation from data retrieval to data analysis including analytical code for both databases (available as supplementary materials on Zenodo) [44], supporting reproducibility, validation, and extension of the findings. Moreover, this study provides methodological guidance for future research aimed at improving understanding of ADWE-related harms in pharmacovigilance data.

However, findings should be interpreted with caution, as there is no definitive evidence of a causal relationship between (withdrawal of) drug exposure and the reported ADWEs. This study used spontaneous reporting databases, which include only voluntarily reported adverse events and are therefore subject to underreporting and reporting bias. In addition, serious events are more likely to be reported than non-serious ones, which may lead to an overrepresentation of more severe outcomes. ADWE incidence cannot be estimated from FDA-FAERS or EMA-EV databases due to incomplete reporting (numerator) and the unknown population at risk (denominator). While spontaneous reports are valuable for pharmacovigilance, they are not sufficient to establish causality, as observed reactions may be related to underlying diseases, unrelated health conditions, or concomitant medications. An additional limitation is the potential overlap of reports between FDA-FAERS and EMA-EV. Duplicate reports were identified and removed within each database according to database-specific procedures; however, deduplication between datasets was not possible due to the absence of common case identifiers. Adverse events may be reported to multiple regulatory authorities; therefore, some cases may be present in both databases. Therefore, overlap between the databases cannot be excluded and may have influenced the reported frequencies of ADWE cases. The number of reports in the databases is influenced by factors such as drug utilisation patterns, awareness among healthcare professionals, and the public. Additionally, reporting may also be affected by the ripple effect (increases of reporting following publicity about drugs in the same class), and the notoriety effect (heightened reporting in response to safety alerts) [5, 84].

There is also no clear distinction between different types of ADWEs (e.g., physiological reactions versus relapse/return of medical conditions). Additionally, there is lack of data on how medications were discontinued (abruptly or tapered, and whether over a short or long period of time). Withdrawal PTs allow us to identify ADWEs in pharmacovigilance data; however, they do not provide comprehensive information about the signs and symptoms of such events. Finally, a high proportion of reports lacked clear documentation of medication indications, age and reporting source, highlighting a significant limitation in pharmacovigilance data quality. Multivariable logistic regression analyses were restricted to complete-case records with available age and sex data, which may introduce selection bias due to missing demographic information and limit the generalisability of the adjusted associations. Accurate and complete documentation is essential for interpreting ADWEs correctly. Previous evidence indicates only 1 out of 8 ADR reports are well documented, highlighting a need for complete and detailed information in reports to support accurate evaluation of causality [85].

### 4.3. Clinical Implications and Future Research

These findings have important implications for clinicians, patients, policymakers, and researchers. The risk of ADWEs should be carefully considered in benefit-risk assessments for individual patients, as similar risks have been observed with different drug classes, including prescription-only, over-the-counter, and controlled drugs. This study emphasises that sex-based variations should be considered during drug discontinuation. Strengthening reporting among males and integrating sex-specific considerations into deprescribing decision-making processes may enhance the effectiveness of ADWE detection and prevention efforts. Extra caution is required for older adults who discontinue long-term treatment and receive high-risk medications, as they may be at higher risk of severe withdrawal reactions with potentially serious consequences. Future research should examine the association between ADWEs and drugs via disproportionality analysis, as well as the variability of ADWE across drug classes and across patient and other characteristics to better identify the strongest signals. Where signals are detected, clinicians should consider the risk of physiological withdrawal reactions on discontinuation when making decisions to initiate such medications. These findings also highlight a lack of explicit guidance on ADWEs within current pharmacovigilance reporting systems, which may contribute to underreporting in routine clinical practice. Additionally, studies are needed to develop standardised criteria for classifying ADWEs and to systematically capture relevant dechallenge and rechallenge data to support evidence-based deprescribing practices and enhance patient safety.

## 5. Conclusion

This study provides novel evidence of reporting patterns and characteristics of ADWEs in pharmacovigilance data. These findings emphasise the need for reporting forms to better accommodate ADWEs (i.e., clarity on terminologies, dechallenge/rechallenge data, causality assessment) to effectively capture ADWE-related data and integrate these into evidence-based deprescribing practices for enhanced patient safety.

## Funding and disclaimer

This research is funded by the European Union through the Marie Skłodowska-Curie Actions (MSCA) programme (Project acronym: HEAD-P; Project number: 101149577) https://cordis.europa.eu/project/id/101149577. Views and opinions expressed are however those of the author(s) only and do not necessarily reflect those of the European Union or the European Research Executive Agency (REA). Neither the European Union nor the granting authority can be held responsible for them.

ER was supported by an NHMRC Investigator Grant (GNT1195460).

## Supporting information

Top 300 PTs in FDA-FAERS database

Top 300 PTs in EMA-EV database

Comparison of structural and variable-level differences between LAERS and FAERS

Variables, PT selection, data cleaning, drug mapping, and reporting sources for FDA-FAERS and EMA-EV (Tables S2.1-S2.7)

Reporter countries, reporting trends, and onset of ADWEs in FDA-FAERS and EMA-EV (Tables S3.1-S3.6).

Information available only in EMA-EV (Tables S4.1-S4.3)

Commonly reported drug classes/drugs and top 200 indications in FDA-FAERS and EMA-EV (Tables S5.1-S5.3)

Chi-square analysis of ADWE seriousness by age and sex in FDA-FAERS and EMA-EV (Tables S6.1-S6.2)

## Data Availability

All data produced in the present study are available in the Supplementary files. The scripts used for data processing and analysis are publicly available on Zenodo at https://zenodo.org/records/18155023.

