## Supplementary material for "Reporting patterns of adverse drug withdrawal events using individual case safety reports in United States and European databases": Comparison of structural and variable-level differences between LAERS and FAERS

**Supplementary file 1:** The structural and variable-level differences between the Legacy Adverse Event Reporting System (LAERS) and the FDA Adverse Event Reporting System (FAERS).

This supplementary table summarises the structural and variable-level differences between the Legacy Adverse Event Reporting System (LAERS; 2004Q1-2012Q3) and the FDA Adverse Event Reporting System (FAERS; 2012Q4-2023Q4). For each data file (DEMO, DRUG, INDI, THER, REAC, OUTC, and RPSR), corresponding variable names, description, and availability periods are presented. These datasets are openly available for scientific use in ASCII (American Standard Code for Information Interchange) and HML (Extensible Markup Language) format. We used ASCII (.txt) files for analysis. LAERS is the earlier version of the database (publicly available since 2004) and was replaced by FAERS on September 10, 2012 as an upgraded system for adverse event reporting.

| **Legacy Adverse Event Reporting System (LAERS)** | | **FDA Adverse Event Reporting System (FAERS)** | |
| --- | --- | --- | --- |
| **2004Q1-2012Q3** | | **2012Q4-2023Q4** | |
| **DEMOGRAPHIC File (DEMO)** | | | |
| **Variable name** | **Description** | **Variable name** | **Description** |
| ISR (7 digits) | Unique number for identifying an AERS report (primary key) (example: 3123456) | ***Primaryid (8 digits)** | Unique number for identifying a FAERS report (primary key) (example: 31234561). |
| Case | Number for identifying an AERS case | ****Caseid** | Number for identifying a FAERS case |
| Caseversion | Not available | ***Caseversion** | Safety report version number. The Initial Case will be version 1; follow-ups to the case will have sequentially incremented version numbers (for example, 2, 3, 4, etc.). |
| I_f_cod | Code for the initial or follow-up status of the report | I_f_cod | Code for the initial or follow-up status of the report |
| Foll_seq | The sequence number of a follow-up report, as reported by the manufacturer. | **Removed** | - |
| Image | Identifier for an AERS report image. Character field consisting of the ISR number, a dash, and a check digit or letter (ex: 3123456-X) | **Removed** | - |
| Event_dt | Date adverse event occurred or began. (YYYYMMDD format) | Event_dt | Available |
| Mfr_dt | Date manufacturer first received initial (or follow-up) information (YYYYMMDD format) | Mfr_dt | Available |
| Fda_dt | Date FDA received report (YYYYMMDD format) | Fda_dt | Available |
| Init_fda_date | Not available | Init_fda_date | Available |
| Rept_cod | Code for the type of report submitted (See table below.)  CODE MEANING_TEXT  ---- ---------------  EXP Expedited (15-Day)  PER Periodic  DIR Direct | Rept_cod | Available |
| Mfr_num | Manufacturer's unique report identifier | Mfr_num | Available |
| Mfr_sndr | Verbatim name of manufacturer sending report | Mfr_sndr | Available |
| Age | Numeric value of patient's age at event. | Age | Available |
| Age_cod | Unit abbreviation for patient's age | Age_cod | Available |
| Age_grp | Not available | ***Age_grp** | Patient age group code as follows, when available: Code meaning_text (N = Neonate; I = Infant; C = Child; T = Adolescent; A = Adult E Elderly). Available during 2014Q3-2023Q4 |
| Gndr_cod | Code for patient's sex (Available during 2004Q1-2012Q3) | Gndr_cod | Code for patient's sex (Available during 2012Q4-2014Q2) |
| - | - | ****Sex** | 2014Q3-Onward Gndr_cod replaced with “Sex” |
| E_sub | Whether (Y/N) this report was submitted under the electronic submissions procedure for manufacturers | E_sub | Available |
| Wt | Numeric value of patient's weight | Wt | Available |
| Wt_cod | Unit abbreviation for patient's weight  CODE MEANING_TEXT  ---- ------------  KG Kilograms  LBS Pounds  GMS Grams | Wt_cod | Available |
| Rept_dt | Date report was sent (YYYYMMDD format) | Rept_dt | Available |
| Occp_cod | Abbreviation for the reporter's type of occupation in the latest version of a case | Occp_cod | Available |
| To_mfr | Whether or not (Y/N) voluntary reporter also notified manufacturer (blank for manufacturer reports). | To_mfr | Available |
| Death_dt | Date patient died (YYYYMMDD format) | Death_dt | Not applicable as removed in 2010Q3-Onwards. This field remains but is no longer populated with data due to privacy concerns |
| Confid | Whether or not (Y/N) voluntary reporter stated that his identity should not be disclosed to the product manufacturer (blank for manufacturer reports). | **Removed** | - |
| *Note*: REPORTER_COUNTRY was not included during 2004Q1 to 2005Q2* | | | |
| Reporter_country | Reporters are asked to give their addresses. This is usually the country the event occurred in. (Available during 2005Q3-2012Q3) | Reporter_country | The country of the reporter in the latest version of a case.  (Available during 2012Q4-2023Q4) |
| Occr_country | Not available | ***Occr_country** | The country where the event occurred (+ New tag added in 2012Q4) |
| **DRUG File (DRUG)** | | | |
| ISR | Available | Primaryid | Available |
| Drugname | Name of medicinal product. If a "Valid Trade Name" is populated for this Case, then DRUGNAME = Valid Trade Name; if not, then DRUGNAME = "Verbatim" name, exactly as entered on the report. Available during 2004Q1-2023Q4 | Drugname | Name of medicinal product. If a "Valid Trade Name" is populated for this case, then DRUGNAME = Valid Trade Name; if not, then DRUGNAME = "Verbatim" name, exactly as entered on the report. Available during 2014Q3-2023Q4 |
| Prod_ai | Not available | ***Prod_ai** | DRUG2014Q3-Onward as "Product Active Ingredient", when available. + New tag added in 2014Q3 extract. |
| Role_cod | Code for drug's reported role in event | Role_cod | Available |
| Drug_seq | Unique number for identifying a drug for an ISR. To link to the THERyyQq.TXT and INDIyyQq. TXT data files, both the ISR number (primary key) and the DRUG_SEQ number (secondary key) are needed. | Drug_seq | Unique number for identifying a drug for a Case. To link to the THERyyQq.TXT and and INDIyyQq data file, both the Case number (primary key) and the DRUG_SEQ number (secondary key) are needed |
| Dechal | Dechallenge code, indicating if reaction abated when drug therapy was stopped | Dechal | Available |
| Rechal | Rechallenge code, indicating if reaction recurred when drug therapy was restarted | Rechal | Available |
| **INDICATION File (INDI)** | | | |
| ISR | Available | Primaryid | Available |
| Drug_seq | Labeled as drug_seq | ****Indi_drug_seq** | Marked with a different label |
| Indi_pt | "Preferred Term" level medical terminology describing the Indication for use. | indi_pt | Available |
| **THERAPY File (THER)** | | | |
| ISR | Available | Primaryid | Available |
| Drug_seq | Labeled as drug_seq | ****Dsg_drug_seq** | Marked with a different label |
| Start_dt | A date therapy was started (or re-started) for drug | Start_dt | Available |
| End_dt | A date therapy was stopped for drug | End_dt | Available |
| Dur | Numeric value of the duration (length) of therapy | Dur | Available |
| Dur_cod | Unit abbreviation for duration of therapy | Dur_cod | Available |
| **REACTION File (REAC)** | | | |
| ISR | Available | Primaryid | Available |
| PT (preferred term) | Available | PT | Available |
| Drug_rec_act | Not available | ***Drug_rec_act** | Drug recur action data - populated with reaction/event information (PT) if/when the event reappears upon readministration of the drug. + New tag added in 2014Q3 extract. |
| **OUTCOME File (OUTC)** | | | |
| ISR | Available | Primaryid | Available |
| Outc_code | Available | Outc_code | Available |
| **REPORTING SOURCE FILE (RPSR)** | | | |
| ISR | Available | Primaryid | Available |
| Rpsr_cod | Available | Rpsr_cod | Available |

*Removed:* indicates variables present in LAERS but no longer populated in FAERS; *Not available:* variables not available during that reporting period; *Bold* variables*: new tag added for variable; *Bold** variable:* same variable marked with different label.
