## Supplementary material for "Reporting patterns of adverse drug withdrawal events using individual case safety reports in United States and European databases": Variables, PT selection, data cleaning, drug mapping, and reporting sources for FDA-FAERS and EMA-EV (Tables S2.1-S2.7)

**Supplementary file 2:** Details of similar variables available in the FDA-FAERS and EMA-EV, details of variables only available in EMA-EV, relevant and non-relevant preferred terms (PTs) containing “withdraw”, data cleaning process, drug cleaning and mapping, variables used and reporting sources/sender type in FDA-FAERS and EMA-EV databases (Tables S2.1, S2.2, S2.3, S2.4, S2.5, S2.6 and S2.7)

**Brief description of FDA-FAERS EMA-EV files**

Each of the FDA-FAERS quarterly data extracts consists of seven files, namely DEMO (demographic), DRUG (drug), INDI (indication), THER (therapy), REAC (reaction), OUTC (outcome), and RPSR (report sources) files *(See supplementary file 1)*. The EMA-EV Level 2A datasets comprise multiple files presented as Excel sheets, including Safety_Report, Patient, Reporter, Drug, Reaction, Literature_Study, Parent, Test, and Diagnosis_Summary. As per the objectives of this study, data were extracted from the safety_report, patient, reporter, drug, and reaction files to obtain the necessary information for analysis. The details of similarities and differences between available variables in both databases are listed in **Table S2.1 and S2.2**.

**Table S2.1:** Details of similar variables available in the FDA-FAERS and EMA-EV databases

| **FDA-FAERS** | | **EMA-EV** | |
| --- | --- | --- | --- |
| **Variable** | **Descriptor** | **Variable** | **Descriptor** |
| **Primaryid** | Unique number for identifying a report | **Evlocalreportnumber** | Unique number assigned to each case |
| **Fda_dt** | Date FDA received case | **First_received_date** | Date on which the report was first received from source |
| **Age** | Numeric value of patient's age at event | **Onset_age** | Age at time of onset of reaction/event (number) |
| **Age_code** | dec = decade;  yr = year;  mon = month;  wk = week;  dy = day;  hr = hour. | **Onset_age_unit** | Age at time of onset of reaction/event (unit) |
| **Age_grp** | n = neonate;  i = infant;  c = child;  t = adolescent  a = adult;  e = elderly | **Patient_age_group_id** | Patient age group =  neonate (Preterm and Term newborns); infant; foetus; child; adolescent; adult; elderly. |
| **Sex** | m = male;  f = female;  unk = unknown | **Patient_sex_id** | m = male;  f = female;  unk = unknown |
| **Occp_cod** | md = physician;  ph = pharmacist;  ot = other healthprofession;  lw = lawyer;  cn = consumer | **Rep_qualification_id** | Qualification (Physician;  pharmacist; consumer or other non-health professional; other health-professional) |
| **Reporter_country** | The country of the reporter | **Country_id** | Reporter’s country code |
| **Prod_ai** | Product active ingredient (drug) | **Recodedproductsccomps** | Recorded product compound (drug) |
| **drugname** | Valid trade name/Verbatim name as entered by the reporter | **-** | - |
| **Role_code** | ps = primary suspect drug;  ss = secondary suspect drug;  c = concomitant;  i = interacting | **characterisation_id** | Characterisation of drug role (Suspect, concomitant, and interacting) |
| **Dechal** | y = positive dechallenge;  n = negative dechallenge;  u = unknown;  d = does not apply | **Action_taken_id** | Action(s) taken with drug: The value ‘1’ (Drug withdrawn), taken together with the ‘Outcome of reaction/event at the time of last observation, describe the dechallenge. |
| **Rechal** | y = positive rechallenge;  n = negative rechallenge;  u = unknown;  d = does not apply | **Reaction_recur_readmin** | Did the reaction recur on re-administration?  (y = positive rechallenge;  n = negative rechallenge;  u = unknown;  n/a = not applicable) |
| **Indi_pt** | Preferred terms for the indication of use, using the medical dictionary for regulatory activities (MedDRA) | **Indication_code-pt** | Preferred terms (MedDRA) |
| **Start_dt** | Date the therapy was started (or re-started) for drug | **Start_date** | Date and time of start of drug |
| **End_dt** | A date therapy was stopped for drug | **End_date** | Date and time of last administration |
| **Dur** | Numeric value of the duration (length) of therapy | **A-drug_admin_duration** | Duration of drug administration (number) |
| **Dur_code** | yr = years;  mon = months;  wk = weeks;  day = days;  hr = hours;  min = minutes;  sec = seconds | **B-drug_admin_duration** | Duration of drug administration (unit) |
| **Pt** | Preferred term for the reaction/event, using the MedDRA | **Reaction_code-pt** | Reaction/event: Preferred term (MedDRA) |
| **Outc_cod** | de = death;  lt = life-threatening;  ho = hospitalization - initial or prolonged;  ds = disability;  ca = congenital anomaly;  ri = required intervention to; prevent permanent impairment/damage;  ot = other serious (important medical event) | **A = is_results_in_death**  **B = is_life_threatening**  **C = is_prolonged_hosp**  **D = is_disabling**  **E = is_congenital_anomaly**  **F = is_other_medically_imp** | A = results in death;  B = life threatening;  C = caused/prolonged hospitalisation;  D = disabling/incapacitating;  E = congenital anomaly/ birth defect;  F = other medically important condition |
| **Rpsr** | Reporting source  (fgn = foreign;  sdy = study;  lit = literature;  csm = consumer;  hp = health professional;  uf = user facility;  cr = company representative;  dt = distributor; oth = other) | **Sender_type** | Sender type: health professional, Other (e.g., distributor, study sponsor) Pharmaceutical company, regional pharmacovigilance centre, regulatory authority |

**Table S2.2:** Details of variables only available in EMA-EV

| **Variables** | **Descriptor** |
| --- | --- |
| **Action_taken_id** | Action(s) taken with drug: Dose reduced/dose not changed/drug withdrawn/not applicable |
| **Assess_result** | Result of assessment: Certain, probable/likely, possible, unlikely |
| **Drug_Assess_Method** | Method of Assessment: Naranjo Algorithm/ Scale, Personal Introspection, Physician Assessment, Summary of Product Characteristics (SmPC), literature, Unknown, World Health Organisation-Uppsala Monitoring Centre (WHO-UMC) causality Assessment etc. |
| **Assess_Result** | Doubtful, likely, probable, highly probable etc. |
| **Drug_Assess_Method** | *EU Method of Assessment |
| **EU_Assess_Result** | EU Result of Assessment (No reasonable possibility or Reasonable possibility) |
| **Start date** | Date of start of reaction/event |
| **End date** | Date of end of reaction/event |
| **Duration** | Duration of reaction/event (number) |
| **Duration unit** | Duration of reaction/event (unit) |
| **Outcome** | Outcome of reaction/event at the time of last observation (Fatal, not recovered/not resolved, recovered/resolved, recovered/resolved with sequelae, recovering/resolving and unknown) |

** In EMA-EV, cases are classified using the EU (European Union) assessment method with two outcomes: ‘reasonable possibility’ and ‘no reasonable possibility’.*

**Table S2.3:** Relevant and non-relevant Preferred Terms (PTs) containing “withdraw” in databases.

| **Relevant PTs** | **Non-Relevant PTs** |
| --- | --- |
| **PTs in FDA-FAERS database** | |
| Withdrawal syndrome | Abnormal withdrawal bleeding |
| Drug withdrawal syndrome | Alcohol withdrawal syndrome |
| Drug withdrawal headache | Anti-androgen withdrawal syndrome |
| Drug withdrawal convulsions | Drug withdrawal maintenance therapy |
| Steroid withdrawal syndrome | Drug withdrawal syndrome neonatal |
| Topical steroid withdrawal reaction | Tobacco withdrawal symptoms |
| Withdrawal catatonia | Withdrawal bleed |
| Withdrawal hypertension | Withdrawal of life support |
| Dopamine agonist withdrawal syndrome | Withdrawal hepatitis |
| - | Thought withdrawal |
| - | Withdrawal bleeding irregular |
| **PTs in EMA-EV database** |  |
| Withdrawal syndrome | Drug withdrawal syndrome neonatal |
| Drug withdrawal syndrome | Alcohol withdrawal syndrome |
| Steroid withdrawal syndrome | Abnormal withdrawal bleeding |
| Drug withdrawal convulsions | Drug withdrawal maintenance therapy |
| Drug withdrawal headache | Thought withdrawal |
| Withdrawal arrhythmia | **-** |
| Withdrawal catatonia | **-** |
| Withdrawal hypertension | **-** |

**Table S2.4:** Data cleaning process (data merging and deduplication)

| **Dataset** | **Primary identifier** | **Deduplication method** |
| --- | --- | --- |
| FDA-FAERS | primaryid | Retain only most recent report based on FDA_DT; remove duplicates using CASE_ID, sex, age, reporter_country, caseversion, and i_f_cod |
| EMA-EV | evlocalreportnumber | Retain only most recent report based on first_received_date; identify duplicates using first_received_date, message_creation_date, onset_age, patient_sex_id, country_id |
| Description of the data cleaning process (data merging and deduplication) | | |
| First, we merged 20-year (2004Q1-2023Q4) FDA (LAERS and FAERS data) and EMA-EV data separately in Stata software. In FDA-FAERS, we mapped "isr" to "primaryid" (as equivalent entities) when merging the LAERS and FAERS datasets. In the FDA-FAERS data, each case is identified by a primary identifier (primaryid) whereas the EMA-EV data use a local report number (evlocalreportnumber) as the unique case identifier ***(see Table S2.1)***. To ensure consistency, we retained only the report with the most recent FDA_DT (date on which the case was received/the case received by FDA) for FAERS, and the most recent first_received_date (date on which the report was first received from source/the report is submitted to EV) for EMA-EV ***(see Table S2.1)***. Additionally, the deduplication procedure was performed to identifies reports with identical values across specific fields. The FDA-FAERS dataset required cleaning to identify and remove potential duplicate reports before analysis. This was included CASE_ID (caseid: number for identifying a FAERS case), sex, age, reporter_country (country of the reporter), caseversion (version of the report), and i_f_cod (indicating whether the report is initial or follow-up) for FDA-FAERS.  In EMA-EV, we also conducted deduplication based on first_received_date, message_creation_date (date of message creation), onset_age, patient_sex_id, and country_id to identify duplicate records. This approach ensured that only a single report was retained for each case. Complete details of the data cleaning process (merging, cleaning and deduplication) with all Stata commands are available on ***Zenodo*** ^[1] *^*.* File details: ***FDA_DataLoop_LAERS_FAERS*** (.do and txt) and ***EMA_DataLoop_EudraVigilance*** (.do and txt) | | |

*[1] *: Khan, Z. and F. Moriarty, Data and materials for "Reporting patterns of adverse drug withdrawal events using individual case safety reports in United States and European databases" (v1.0). Zenodo. 2026: https://zenodo.org/records/18155023.*

**Table S2.5:** Description drug cleaning and mapping

| **Dataset** | **Drug Variables** | **Availability** | **Cleaning / Mapping Steps** |
| --- | --- | --- | --- |
| FDA-FAERS | **drugname** (a mixture of entries, including valid trade names and verbatim text as reported, which contain pharmaceutical company names, dosage forms, or other irrelevant text) | 2004Q1 to 2023Q4 | 1. Removed irrelevant terms from drugname (pharma company names, dosage forms, etc.) 2. Mapped brand names to generic active ingredients using RxNorm via RxNav and Drugs@FDA 3. Used cleaned drugname to fill missing prod_ai for 2004Q1-2014Q2 4. Final cleaned prod_ai used in main analysis |
|  | **prod_ai** (product active ingredient) | 2014Q3-2023Q4 |  |
| EMA-EV | recodedproductsccomps | 2004Q1 to 2023Q4 | Already cleaned variable provided by the EMA |
| **Description of the drug cleaning and mapping** | | | |
| In the FDA-FAERS drug files, two variables include drugname (a mixture of entries, including valid trade names and verbatim text as reported, which contain pharmaceutical company names, dosage forms, or other irrelevant text) and prod_ai (product active ingredient). The drugname variable is available from 2004Q1 to 2023Q4, while prod_ai was introduced as a new tag in the 2014Q3 extract. To ensure consistency, we extensively cleaned drugname (removal of irrelevant terms) and mapped brand names to their generic equivalents. The cleaned drugname variable values were then used to replace missing prod_ai entries for the earlier years (2004Q1–2014Q2). We then used a final cleaned prod_ai variable in the main analysis. We used RxNorm via RxNav and Drugs@FDA, to map brand names to their corresponding generic active ingredients. RxNorm is a standardised nomenclature developed by the US National Library of Medicine (NLM) that links across widely used drug vocabularies (e.g., First Databank, Micromedex, Multum, Gold Standard Drug Database). It also incorporates the US Pharmacopeia, which provides a comprehensive listing of all active pharmaceutical ingredients (APIs). The detailed Stata script for drug cleaning and mapping for FDA data is available on ***Zenodo*** ^[1] *^ ***(FDA_DrugCleaning_and_Mapping*** (.do and txt) .  In the EMA-EV, we used the already cleaned drug name variable “RecodedProductsComps” (Active Substance, High Level) provided by EMA, which represents the standardised, non-proprietary name of the APIs. | | | |

*[1] *: Khan, Z. and F. Moriarty, Data and materials for "Reporting patterns of adverse drug withdrawal events using individual case safety reports in United States and European databases" (v1.0). Zenodo. 2026: https://zenodo.org/records/18155023.*

**Table S2.6:** Variables used for analysis and their availability in both databases

| **Variable** | **Description and availability of variables in databases** |
| --- | --- |
| **Patient characteristics** | |
| **Age group** | **FDA-FAERS:** Neonate; Infant; Child; Adolescent; Adult; Elderly**.**  **EMA-EV:** Neonate (preterm and term newborns); Infant; Foetus; Child; Adolescent; Adult; Elderly.  **Harmonisation:** *Neonate to adolescent grouped* *as Paediatric (0-17 years); adults 18-64 years; elderly ≥65 years (Older adults).* |
| **Sex** | **Both databases:** Male; Female; Not specified |
| **Reporter information** | |
| **Reporter type** | **Both databases:** Consumer or other non-health professional (e.g., lawyer); Physician; Pharmacist; Other healthcare professional; Not specified |
| **Reporting source / Sender type** | **FDA-FAERS:** Consumer; Company representative; Foreign; Literature; User facility**.**  **EMA-EV:** Pharmaceutical company; Regulatory authority; Regional pharmacovigilance centre.  **Both databases:** Health professional organisation; Other (e.g., distributor, study sponsor); Not specified. |
| **Reporter country** | **Both databases:** Country name of primary reporter |
| **Year of reporting** | **Both databases:** 2004 to 2023 |
| **Drug information** | |
| **Drug name** | **Both databases:** Standardised, non-proprietary name of the active product ingredients (APIs). |
| **Drug class** | **Both databases:** WHO-Anatomical Therapeutic Chemical (ATC) Therapeutic drug class |
| **Role** | **FDA-FAERS:** Primary suspect; secondary suspect.  **EMA-EV:** Suspect. |
| **Indication** | **Both databases:** Indication for drug use |
| **ADWE characteristics** | |
| **ADWE (Preferred Term)** | **Both databases:** MedDRA Preferred Term (PT) |
| **Seriousness criteria** | **Both databases:** Death; Life-threatening; Hospitalisation; Disability; Congenital anomaly/birth defect; Other medically important condition.  **FDA-FAERS:** Required intervention to prevent permanent impairment/damage. |
| **Time to onset** | **FDA-FAERS:** Event date − drug start date.  **EMA-EV:** Reaction start date − drug start date. |
| **Dechallenge** | **Both databases:** Unknown (dechallenge was done, outcome unknown); Not applicable/does not apply  **FDA-FAERS:** Yes (dechallenge was done, reaction resolved); No (dechallenge was done, reaction did not resolve).  **EMA-EV:** Drug withdrawn; Dose not changed; Dose reduced; Dose increased; Outcome as recovered/resolved |
| **Rechallenge** | **Both databases:**  **Yes - unknown** (rechallenge was done, outcome unknown);  **No - n/a** (no rechallenge was done, recurrence is not applicable);  **Yes - no** (rechallenge was done, reaction did not recur);  **Yes - yes** (rechallenge was done, reaction recurred) |
| **EMA-only variables** | |
| **Duration of ADWE** | Reaction start date - reaction end date |
| **Action taken with drug** | Drug withdrawn; Dose reduced; Dose increased; Dose not changed |
| **Causality** *(defined as an estimation of the supposed causal relationship between a drug treatment and the occurrence of an adverse event)* **assessment method** | Naranjo algorithm/scale; WHO–Uppsala Monitoring Centre assessment; Physician assessment; Personal introspection; Literature-based evaluation |
| **EU-specific causality outcome** | Reasonable possibility; No reasonable possibility |
| **Outcome at last observation** | Fatal; Not recovered/not resolved; Recovered/resolved; Recovering/resolving; Unknown |

**Table S2.7:** Reporting sources/sender type in FDA-FAERS and EMA-EV database

| ****Reporting sources/sender type** | **FDA-FAERS *n* (%)** | **EMA-EV *n* (%)** |
| --- | --- | --- |
| Consumer | 7,275 (5) | **-** |
| Health professional | 6,989 (4.8) | 2143 (16.5) |
| Other (e.g., distributor, study sponsor) | 3,104 (2.1) | 266 (2) |
| Foreign (*n*=1,902), company representative (*n*=1,843), Literature (*n*=835) and user facility (*n*=15) | 4595 (3.1) | - |
| Pharmaceutical company | - | *****3198 (24.6) |
| Regulatory authority | - | *****7103 (54.7) |
| Regional pharmacovigilance centre | - | *77 (0.6) |
| Not specified | 123,551 (84.9) | 200 (1.5) |
| ***Total*** | ***145,514*** | ***12,987*** |

******* *Only available in EMA -EV data, **Reporting sources/sender type is reported as described in both datasets (see Supplementary File 2-Table S2.1). In FDA data, the reporting sources was not specified for 84.9% of reports; the primary reporting sources were consumers (5.8%) and health professional (4.8%). In EMA data, regulatory authorities (54.7%) and pharmaceutical companies (24.6%) were the leading sources of ADWE reports.*
