## Supplementary material for "Reporting patterns of adverse drug withdrawal events using individual case safety reports in United States and European databases": Reporter countries, reporting trends, and onset of ADWEs in FDA-FAERS and EMA-EV (Tables S3.1-S3.6).

**Supplementary file 3:** Reporter countries, reporting trends over time and onset of ADWEs in FDA-FAERS and EMA-EV database (Tables S3.1, S3.2, S3.3, S3.4, S3.5 and S3.6)

**Table S3.1:** Details of reporter countries in FDA-FAERS database

| **FDA-FAERS database** | | |
| --- | --- | --- |
| **Reporter country** | **Frequency** | **Percentage** |
| United States | 113,523 | 78.02 |
| United Kingdom | 8,343 | 5.73 |
| **Country not specified** | 8,151 | 5.60 |
| Canada | 2,883 | 1.98 |
| France | 2,520 | 1.73 |
| Germany | 1,890 | 1.30 |
| Japan | 1,303 | 0.90 |
| Australia | 952 | 0.65 |
| Netherlands | 610 | 0.42 |
| Italy | 487 | 0.33 |
| Brazil | 458 | 0.31 |
| Denmark | 443 | 0.30 |
| Spain | 398 | 0.27 |
| Sweden | 367 | 0.25 |
| Switzerland | 340 | 0.23 |
| India | 317 | 0.22 |
| Ireland | 232 | 0.16 |
| Turkey | 182 | 0.13 |
| Poland | 169 | 0.12 |
| Finland | 167 | 0.11 |
| Norway | 148 | 0.10 |
| Portugal | 148 | 0.10 |
| Taiwan | 130 | 0.09 |
| China | 126 | 0.09 |
| Austria | 120 | 0.08 |
| Belgium | 114 | 0.08 |
| Czechia | 103 | 0.07 |
| New Zealand | 82 | 0.06 |
| Iran | 79 | 0.05 |
| Israel | 50 | 0.03 |
| Greece | 48 | 0.03 |
| South Africa | 47 | 0.03 |
| Argentina | 45 | 0.03 |
| Korea, Republic of (South Korea) | 44 | 0.03 |
| Mexico | 40 | 0.03 |
| Hungary | 39 | 0.03 |
| Russia | 34 | 0.02 |
| Croatia | 30 | 0.02 |
| Colombia | 29 | 0.02 |
| Malaysia | 23 | 0.02 |
| Singapore | 18 | 0.01 |
| Puerto Rico | 17 | 0.01 |
| Slovenia | 16 | 0.01 |
| Egypt | 15 | 0.01 |
| Chile | 14 | 0.01 |
| Tunisia | 14 | 0.01 |
| Romania | 13 | 0.01 |
| Thailand | 12 | 0.01 |
| Venezuela | 11 | 0.01 |
| **Others (≤10)** | 92 | 0.09 |
| **Total** | 145,514 | 100.00 |

**Table S3.2:** Details of reporter countries in EMA-EV database

| **EMA-EV database** | | |
| --- | --- | --- |
| **Reporter Country** | **Frequency** | **Percentage (%)** |
| France | 3,624 | 27.9% |
| Germany | 2,674 | 20.6% |
| United Kingdom | 2,439 | 18.8% |
| Netherlands | 1,035 | 8.0% |
| Sweden | 599 | 4.6% |
| Spain | 387 | 3.0% |
| Italy | 385 | 2.96% |
| Denmark | 273 | 2.1% |
| Finland | 229 | 1.76% |
| Norway | 213 | 1.64% |
| Belgium | 208 | 1.6% |
| Ireland | 207 | 1.6% |
| Poland | 168 | 1.3% |
| Austria | 149 | 1.15% |
| Greece | 79 | 0.61% |
| Portugal | 75 | 0.58% |
| Czech Republic | 67 | 0.52% |
| Hungary | 45 | 0.35% |
| Croatia | 34 | 0.26% |
| Slovenia | 23 | 0.18% |
| Romania | 17 | 0.13% |
| Iceland | 11 | 0.08% |
| Slovakia | 11 | 0.08% |
| Others (≤10) | 35 | 0.27% |
| Total | 12,987 | 100.00% |

**Table S3.3:** ADWE reporting trends over time (2004–2023)

| **Variables** | **FDA *n* (%)** | **EMA *n* (%)** |
| --- | --- | --- |
| 2004 | 4490 (3.1) | 106 (0.8) |
| 2005 | 4910 (3.4) | 153 (1.2) |
| 2006 | 2597 (1.8) | 280 (2.1) |
| 2007 | 2738 (1.9) | 459 (3.5) |
| 2008 | 3757 (2.6) | 402 (3.1) |
| 2009 | 2689 (1.8) | 447 (3.4) |
| 2010 | 5043 (3.5) | 472 (3.6) |
| 2011 | 3962 (2.7) | 468 (3.6) |
| 2012 | 4242 (2.92) | 521 (4.0) |
| 2013 | 4412 (3) | 573 (4.4) |
| 2014 | 4270 (2.93) | 663 (5.1) |
| 2015 | 8063 (5.5) | 624 (4.8) |
| 2016 | 6725 (4.6) | 744 (5.7) |
| 2017 | 8807 (6) | 841 (6.5) |
| 2018 | 6995 (4.8) | 1256 (9.6) |
| 2019 | 7388 (5.1) | 1221 (9.4) |
| 2020 | 9960 (6.8) | 1323 (10.2) |
| 2021 | 24316 (16.7) | 898 (6.9) |
| 2022 | 22183 (15.2) | 809 (6.2) |
| 2023 | 7967 (5.5) | 727 (5.6) |
| **2004-2023** | **145514 (100)** | **12987 (100)** |

**Table S3.4:** Overall Onset of ADWEs (FDA-FAERS=10,996 vs EMA-EV=3,701)

| **Onset of reactions** | **FDA *n* (%)** | **FDA *cum* %** | **EMA *n* (%)** | **EMA *cum* %** |
| --- | --- | --- | --- | --- |
| 1 day or less | 3141 (28.6) | 28.6 | 576 (15.6) | 15.6 |
| 2-3 days | 386 (3.5) | 32.1 | 79 (2.1) | 17.7 |
| 4-7 days | 452 (4.1) | 36.2 | 131 (3.5) | 21.2 |
| 8-14 days | 421 (3.8) | 40.0 | 125 (3.4) | 24.6 |
| 15-30 days | 737 (6.7) | 46.7 | 215 (5.8) | 30.4 |
| 1-3 months | 1246 (11.3) | 58.0 | 402 (10.9) | 41.3 |
| 3-6 months | 980 (8.9) | 66.9 | 371 (10.0) | 51.3 |
| 6-12 months | 1079 (9.8) | 76.7 | 511 (13.8) | 65.1 |
| Over 1 year | 2554 (23.2) | 100 | 1291 (34.9) | 100 |

*Cum %=cumulative %*

**Table S3.5:** Onset of ADWEs in FDA-FAERS database (*n*=10,996)

| **PT** | **≤1 d** | **2–3 d** | **4–7 d** | **8–14 d** | **15–30 d** | **1–3 m** | **3–6 m** | **6–12 m** | **>1 y** | **Total** |
| --- | --- | --- | --- | --- | --- | --- | --- | --- | --- | --- |
| Drug withdrawal syndrome | 1,824 (30.9) | 199 (3.4) | 235  (4) | 241 (4.1) | 396  (6.7) | 683  (11.6) | 563  (9.5) | 578  (9.8) | 1,186 (20.1) | **5,905** |
| Withdrawal syndrome | 1,246 (27.1) | 172 (3.7) | 201 (4.4) | 168 (3.6) | 315  (6.8) | 490  (10.6) | 386  (8.4) | 449  (9.8) | 1,175 (25.5) | **4,602** |
| Steroid withdrawal syndrome | 19  (9.8) | 2  (1) | 3  (1.5) | 4  (2.1) | 14  (7.2) | 28  (14.4) | 10  (5.2) | 23  (11.9) | 91  (46.9) | **194** |
| Drug withdrawal convulsions | 27  (14.2) | 8  (4.2) | 9  (4.7) | 5  (2.6) | 8  (4.2) | 26  (13.7) | 15  (7.9) | 24  (12.6) | 68  (35.8) | **190** |
| Drug withdrawal headache | 13  (27.7) | 2  (4.3) | 1  (2.1) | 1  (2.1) | 4  (8.5) | 7  (14.9) | 2  (4.3) | 2  (4.3) | 15  (31.9) | **47** |
| Topical steroid withdrawal | 1  (3.3) | 0  (0) | 2  (6.7) | 1  (3.3) | 0  (0) | 9  (30) | 3  (10) | 1  (3.3) | 13  (43.3) | **30** |
| Withdrawal hypertension | 10  (38.5) | 3 (11.5) | 1  (3.8) | 1  (3.8) | 0  (0) | 3  (11.5) | 1  (3.8) | 1  (3.8) | 6  (23.1) | **26** |
| Withdrawal arrhythmia | 1  (50) | 0  (0) | 0  (0) | 0  (0) | 0  (0) | 0  (0) | 0  (0) | 1  (50) | 0  (0) | **2** |
| **Total n (%)** | 3,141 (28.6) | 386 (3.5) | 452 (4.1) | 421 (3.8) | 737  (6.7) | 1,246 (11.3) | 980  (8.9) | 1,079  (9.8) | 2,554 (23.2) | **10,996** |

*PT = preferred term; d = day; m = month; y = year*

**Table S3.6:** Onset of ADWEs in EMA-EV database (*n*=3,701)

| **PT** | **≤1 d** | **2–3 d** | **4–7 d** | **8–14 d** | **15–30 d** | **1–3 m** | **3–6 m** | **6–12 m** | **>1 y** | **Total** |
| --- | --- | --- | --- | --- | --- | --- | --- | --- | --- | --- |
| Withdrawal syndrome | 377  (15) | 56 (2.2) | 81 (3.2) | 72  (2.9) | 141 (5.6) | 274 (10.9) | 239  (9.5) | 365 (14.6) | 903  (36) | **2,508** |
| Drug withdrawal syndrome | 169  (16.8) | 18 (1.8) | 41 (4.1) | 42  (4.2) | 62  (6.2) | 106  (10.6) | 114  (11.3) | 123  (12.2) | 330  (32.8) | **1,005** |
| Drug withdrawal headache | 10  (16.7) | 1  (1.7) | 2 (3.3) | 2  (3.3) | 1  (1.7) | 12  (20) | 5  (8.3) | 8  (13.3) | 19  (31.7) | **60** |
| Steroid withdrawal syndrome | 3  (5.5) | 2  (3.6) | 3  (5.5) | 4  (7.3) | 5  (9.1) | 4  (7.3) | 6  (10.9) | 8  (14.5) | 20  (36.4) | **55** |
| Drug withdrawal convulsions | 9  (18.8) | 1  (2.1) | 1  (2.1) | 3  (6.3) | 5  (10.4) | 4  (8.3) | 7  (14.6) | 4  (8.3) | 14  (29.2) | **48** |
| Withdrawal hypertension | 7  (36.8) | 1 (5.3) | 3 (15.8) | 2  (10.5) | 1  (5.3) | 0  (0) | 0  (0) | 2  (10.5) | 3  (15.8) | **19** |
| Withdrawal arrhythmia | 1  (33.3) | 0  (0) | 0  (0) | 0  (0) | 0  (0) | 1  (33.3) | 0  (0) | 0  (0) | 1  (33.3) | **3** |
| Withdrawal catatonia | 0  (0) | 0  (0) | 0  (0) | 0  (0) | 0  (0) | 1  (33.3) | 0  (0) | 1  (33.3) | 1  (33.3) | **3** |
| **Total n (%)** | 576  (15.6) | 79 (2.1) | 131 (3.5) | 125  (3.4) | 215 (5.8) | 402 (10.9) | 371  (10) | 511 (13.8) | 1,291 (34.9) | **3,701** |

*PT = preferred term; d = day; m = month; y = year*
