## Supplementary material for "Reporting patterns of adverse drug withdrawal events using individual case safety reports in United States and European databases": Information available only in EMA-EV (Tables S4.1-S4.3)

**Supplementary file 4. Information only available in EMA-EV**

**(Tables S4.1, S4.2 an S4.3)**

**Table S4.1:** Information only available in EMA-EV

| **Outcome of ADWEs** | **EMA-EV *n* (%)** |
| --- | --- |
| Unknown | 5,109 (38.8) |
| Recovered/resolved | 4,274 (32.4) |
| Not recovered/not resolved | 2,266 (17.2) |
| Recovering/resolving | 1,377 (10.4) |
| Recovered/resolved with sequelae | 109 (0.8) |
| Fatal | 41 (0.3) |
| ***Total cases*** | ***13,176*** |
| **Methods used for causality assessment** | |
| Clinical judgment or global introspection | 8,785 (70) |
| WHO-UMC method | 892 (7.1) |
| French imputability method | 851 (6.8) |
| Naranjo algorithm | 459 (3.6) |
| Karch-Lasagna algorithm | 186 (1.5) |
| Manual/personal assessment | 164 (1.3) |
| Eudravigilance clinical trial module (EVCTM) | 116 (0.92) |
| Literature | 4 (0.03) |
| EU method of assessment | 2 (0.02) |
| Summary of product characteristics (SmPC) | 1 (0.01) |
| SmPC and literature | 1 (0.01) |
| Unknown | 1,095 (8.7) |
| ***Total cases*** | ***12,556*** |
| **Results of ADWE causality assessment (including all cases)** | |
| Possible | 5,393 (32.9) |
| Probable/likely | 2,859 (17.4) |
| Unassessable/unclassifiable | 1,834 (11.2) |
| Suspect | 789 (4.8) |
| Unlikely | 367 (2.2) |
| Conditional/unclassified | 335 (2) |
| Certain/definite | 134 (0.8) |
| Doubtful | 131 (0.8) |
| Unknown | 4,554 (27.8) |
| ***Total cases*** | ***16,396*** |
| **Results of ADWE causality assessment (excluding the unassessable/unclassifiable and unknown cases)** | |
| Possible | 5,393 (53.9) |
| Probable/likely | 2,859 (28.6) |
| Suspect | 789 (7.9) |
| Unlikely | 367 (3.7) |
| Conditional/unclassified | 335 (3.3) |
| Certain/definite | 134 (1.3) |
| Doubtful | 131 (1.3) |
| ***Total cases*** | **10,008** |
| **EU-specific assessment outcomes** | |
| Reasonable possibility | 1294 (80.1) |
| No reasonable possibility | 321 (19.9) |
| ***Total cases*** | ***1,615*** |

**Explanations: Methods used for causality assessment**

1. Clinical judgment or global introspection is a subjective assessment of ADR causality conducted by a clinical expert, based on personal knowledge and experience without using a standardised tool) (1).
2. WHO-UMC method is a structured tool that categorises causality into different levels/certain, probable, possible, unlikely, conditional and unassessable based on the strength of the association between drug and the adverse event (2).
3. French Imputability is an algorithmic tool based on eight criteria divided into three groups: chronology, semiology, and bibliographic data. It calculates an intrinsic causality score by combining a chronological score (time to onset, dechallenge, rechallenge) and a semiological score (alternative causes, characteristic of reaction pattern, risk factors, and test results). An additional bibliographic criterion assesses the drug’s known potential to cause the reaction to derive an extrinsic score based on scientific literature (3).
4. Naranjo is an ADR probability scale consists of 10 questions with associated scores for calculating the likelihood of a causal relationship. A total score ranges from -4 to +13; the reaction is considered definite if the score is 9 or higher, probable if 5 to 8, possible if 1 to 4, and doubtful if 0 or less) (4).
5. Karch-Lasagna algorithm consists of three sections including identification of potential drug-related events, assessment of the link between the suspected drug-event and cause of the drug-related event. Each section contains a series of closed (yes/no) questions. The combination of responses is used to establish the causal relationship and the reaction is then classified as related, probable, possible, conditional, or unrelated) (5).
6. The EudraVigilance Clinical Trial Module: EVCTM is a part of the EMA safety database system that collects and manages electronic reports from clinical trial sponsors about serious and unexpected adverse reactions (SUSARs) that occur during interventional clinical studies in the EEA (6).

**Results of ADWEs causality assessment**

i. Possible: The reaction may be caused by the drug, but other causes such as underlying disease, concurrent medications cannot be excluded and the effect of drug withdrawal may be unclear.
ii. Probable/Likely: The reaction is likely caused by the drug, unlikely to be attributed to other causes (disease or other drugs), and symptoms improve if the drug is stopped
iii. Unassessable/Unclassifiable: Insufficient information to assess causality
iv. Suspect: A potential link to the drug is suspected, but supporting evidence is limited, and alternative explanations cannot be ruled out.
v. Unlikely: The drug is unlikely to be the cause and other causes provide more plausible explanations.
vi. Conditional/Unclassified: Data are incomplete or under review and more data needed for proper assessment.
vii. Certain/Definite: Clear evidence shows the drug caused the reaction, other causes are excluded and withdrawal or rechallenge confirms the event
viii. Doubtful: unlikely and inconsistent with known drug effects
ix. Unknown: Causality was not documented (2,7).

**Table S4.2:** Overall duration of ADWEs in EMA-EV database (*n*=1403)

| **Duration** | **Frequency** | **Percentage (%)** | **Cumulative %** |
| --- | --- | --- | --- |
| 1 day or less | 303 | 21.6 | 21.6 |
| 2-3 days | 322 | 22.9 | 44.5 |
| 4-7 days | 268 | 19.1 | 63.6 |
| 8-14 days | 213 | 15.2 | 78.8 |
| 15-30 days | 146 | 10.4 | 89.2 |
| 1-3 months | 83 | 5.9 | 95.2 |
| 3-6 months | 22 | 1.6 | 96.7 |
| 6-12 months | 26 | 1.8 | 98.6 |
| Over 1 year | 20 | 1.4 | 100 |

**Table S4.3:** Duration of relevant ADWE PTs in EMA-EV database

| **ADWEs** | **≤1 d** | **2–3 d** | **4–7 d** | **8–14 d** | **15–30 d** | **1–3 m** | **3–6 m** | **6–12 m** | **>1 y** | **Total (n)** |
| --- | --- | --- | --- | --- | --- | --- | --- | --- | --- | --- |
| Withdrawal syndrome | 214 (22.1) | 215  (22.2) | 189  (19.5) | 153  (15.8) | 99  (10.2) | 56  (5.8) | 16  (1.7) | 16  (1.7) | 9  (0.9) | **967** |
| Drug withdrawal syndrome | 64  (17.2) | 95  (25.5) | 71  (19.1) | 56  (15.1) | 44  (11.8) | 21  (5.6) | 5  (1.3) | 7  (1.9) | 9  (2.4) | **372** |
| Drug withdrawal convulsions | 23  (65.7) | 7  (20) | 3  (8.6) | 0  (0) | 0  (0) | 0  (0) | 0  (0) | 2  (5.7) | 0  (0) | **35** |
| Drug withdrawal headache | 0  (0) | 3  (30) | 2  (20) | 1  (10) | 0  (0) | 3  (30) | 1  (10) | 0  (0) | 0  (0) | **10** |
| Steroid withdrawal syndrome | 1  (11.1) | 0  (0) | 0  (0) | 1  (11.1) | 1  (11.1) | 3  (33.3) | 0  (0) | 1  (11.1) | 2  (22.2) | **9** |
| Withdrawal hypertension | 1  (20) | 2  (40) | 0  (0) | 1  (20) | 1  (20) | 0  (0) | 0  (0) | 0  (0) | 0  (0) | **5** |
| Withdrawal catatonia | 0  (0) | 0  (0) | 2  (66.7) | 1  (33.3) | 0  (0) | 0  (0) | 0  (0) | 0  (0) | 0  (0) | **3** |
| Withdrawal arrhythmia | 0  (0) | 0  (0) | 1  (50) | 0  (0) | 1  (50) | 0  (0) | 0  (0) | 0  (0) | 0  (0) | **2** |
| **Total n (%)** | 303 (21.6) | 322  (22.9) | 268  (19.1) | 213  (15.2) | 146  (10.4) | 83  (5.9) | 22  (1.6) | 26  (1.9) | 20  (1.4) | **1403**  **(100)** |

*PT = preferred term; d = day; m = month; y = year*
