## Supplementary material for "Reporting patterns of adverse drug withdrawal events using individual case safety reports in United States and European databases": Commonly reported drug classes/drugs and top 200 indications in FDA-FAERS and EMA-EV (Tables S5.1-S5.3)

**Supplementary file 5**: Details of commonly reported drug classes/drugs and top 200 indication in FDA-FAERS and EMA-EV databases (Table S5.1, S5.2 and S5.3)

****Note:***  All reported drugs were extracted from each database and individual drugs were grouped into their respective drug classes. Percentages for drug classes were calculated using the total number of reported drugs as the denominator (FDA-FAERS: *n* = 145,552; EMA-EV: *n* = 17,525) (e.g., ***Opioids,*** FDA-FAERS: 63,168/145,552 = 43.4%; EMA-EV: 3,990/17,525 = 22.7%). Frequencies and percentages for individual drugs were calculated using the total number of reports within the corresponding drug class (e.g., ***Oxycodone*,** FDA-FAERS: 18,824/63,168 = 29.8%; ***Buprenorphine***, EMA-EV: 759/3,990 = 19%). Small cell counts (<5, or <10 where required) were suppressed for confidentiality.

**Table S5.1:** Details of drug classes and individual drugs in FDA-FAERS and EMA-EV databases

| **Drugs** | **FDA-FAERS**  ***n* (%)** | **Drugs** | **EMA-EV**  ***n* (%)** |
| --- | --- | --- | --- |
| ***Opioids n (%)*** | ***63168 (43.4)*** | ***Opioids*** | ***3990 (22.7)*** |
| Oxycodone | 18824 (29.8) | Buprenorphine | 759 (19) |
| Hydrocodone acetaminophen | 7878 (12.5) | Tramadol | 669 (16.8) |
| Morphine | 7656 (12.1) | Fentanyl | 567 (14.2) |
| Fentanyl | 6538 (10.4) | Methadone | 437 (11) |
| Hydromorphone | 4226 (6.7) | Morphine | 371 (9.3) |
| Buprenorphine naloxone | 3664 (5.8) | Oxycodone | 344 (8.6) |
| Oxycodone acetaminophen | 3405 (5.4) | Diamorphine | 181 (4.5) |
| Buprenorphine | 3403 (5.4) | Codeine acetaminophen | 138 (3.5) |
| Tramadol | 2386 (3.8) | Oxycodone naloxone | 125 (3.1) |
| Methadone | 1878 (3.0) | Tapentadol | 87 (2.2) |
| Oxymorphone | 1144 (1.8) | Buprenorphine naloxone | 78 (2) |
| Diamorphine | 717 (1.1) | Hydromorphone | 77 (1.9) |
| ***Antidepressants n (%)*** | ***32092 (******22)*** | ***Antidepressants*** | ***3231 (18.4)*** |
| Duloxetine | 10274 (32) | Venlafaxine | 838 (25.9) |
| Paroxetine | 9585 (29.9) | Paroxetine | 489 (15.1) |
| Venlafaxine | 5221 (16.3) | Duloxetine | 374 (11.6) |
| Desvenlafaxine | 2077 (6.5) | Sertraline | 328 (10.2) |
| Sertraline | 1454 (4.5) | Escitalopram | 285 (8.8) |
| Escitalopram | 610 (1.9) | Citalopram | 218 (6.7) |
| Citalopram | 568 (1.8) | Mirtazapine | 172 (5.3) |
| Fluoxetine | 539 (1.7) | Fluoxetine | 114 (3.5) |
| Bupropion | 486 (1.5) | Amitriptyline | 98 (3) |
| Mirtazapine | 423 (1.3) | Clomipramine | 52 (1.6) |
| - | - | Bupropion | 39 (1.2) |
| - | - | Tianeptine | 34 (1.1) |
| ***Gabapentinoids n (%)*** | ***9744 (6.7)*** | ***Gabapentinoids*** | ***1058 (6)*** |
| Pregabalin | 8463 (86.9) | Pregabalin | 927 (87.6) |
| Gabapentin | 1281 (13.1) | Gabapentin | 131 (12.4) |
| ***Hypnotics and sedatives n (%)*** | ***6436 (4.4)*** | ***Hypnotics and sedatives*** | ***2078 (11.8)*** |
| Alprazolam | 1565 (24.3) | Alprazolam | 320 (15.4) |
| Clonazepam | 1490 (23.2) | Lorazepam | 267 (12.8) |
| Lorazepam | 843 (13.1) | Oxazepam | 256 (12.3) |
| Zolpidem | 653 (10.1) | Diazepam | 254 (12.2) |
| Diazepam | 615 (9.6) | Zolpidem | 226 (10.9) |
| Clobazam | 273 (4.2) | Bromazepam | 194 (9.3) |
| Scopolamine | 159 (2.5) | Zopiclone | 185 (8.9) |
| Zopiclone | 124 (1.9) | Clonazepam | 163 (7.8) |
| Oxazepam | 117 (1.8) | Lormetazepam | 42 (2) |
| Dexmedetomidine | 107 (1.7) | Flunitrazepam | 25 (1.2) |
| Temazepam | 91 (1.4) | Temazepam | 23 (1.1) |
| ***Muscle relaxants n (%)*** | ***4512 (3.1)*** | ***Muscle relaxants*** | ***284 (1.6)*** |
| Baclofen | 4305 (95.4) | Baclofen | 273 (96.1) |
| Carisoprodol | 98 (2.2) | Tizanidine | 6 (2.1) |
| Tizanidine | 87 (1.9) | Botulinum toxin | 2 (0.7) |
| Botulinum toxin | 13 (0.3) | Methocarbamol | 1 (0.4) |
| Methocarbamol | 4 (0.1) | Orphenadrine | 1 (0.4) |
| Dantrolene | 3 (0.1) | Thiocolchicoside | 1 (0.4) |
| ***Antipsychotics n (%)*** | ***4471 (3)*** | ***Antipsychotics*** | ***905 (5.1)*** |
| Quetiapine | 1519 (34) | Quetiapine | 251 (27.7) |
| Olanzapine | 562 (12.6) | Olanzapine | 135 (14.9) |
| Aripiprazole | 496 (11.1) | Clozapine | 117 (12.9) |
| Clozapine | 442 (9.9) | Risperidone | 105 (11.6) |
| Risperidone | 361 (8.1) | Aripiprazole | 67 (7.4) |
| Ziprasidone | 260 (5.8) | Lithium | 41 (4.5) |
| Lurasidone | 202 (4.5) | Haloperidol | 33 (3.6) |
| Lithium | 135 (3) | Cyamemazine | 31 (3.4) |
| Haloperidol | 116 (2.6) | Flupentixol | 19 (2.1) |
| Paliperidone | 85 (1.9) | Ziprasidone | 18 (2) |
| Brexpiprazole | 68 (1.5) | Zuclopenthixol | 17 (1.9) |
| - | - | Sulpiride | 16 (1.8) |
| - | - | Chlorpromazine | 12 (1.3) |
| - | - | Loxapine | 12 (1.3) |
| - | - | Pimozide | 9 (1%) |
| ***Drugs used in addictive disorders n (%)*** | ***2335 (1.6)*** | ***Drugs used in addictive disorders*** | ***362 (2)*** |
| Varenicline | 893 (38.2) | Varenicline | 159 (43.9) |
| Naltrexone | 877 (37.6) | Nicotine | 63 (17.4) |
| Nicotine | 530 (22.7) | Nalmefene | 59 (16.3) |
| Nalmefene | 15 (0.6) | levomethadone | 47 (13) |
| Lofexidine | 12 (0.5) | Naltrexone | 31 (8.6) |
| Acamprosate | <10 (0.4) | Disulfiram | <5 (1.1) |
| ***Antiepileptics n (%)*** | ***2052 (1.4)*** | ***Antiepileptics*** | ***367 (2.1)*** |
| Vigabatrin | 528 (25.7) | Valproic acid | 92 (25.1) |
| Lamotrigine | 386 (18.8) | Lamotrigine | 69 (18.8) |
| Valproic acid | 296 (14.4) | Carbamazepine | 50 (13.6) |
| Topiramate | 195 (9.5) | Levetiracetam | 48 (13.1) |
| Levetiracetam | 191 (9.3) | Topiramate | 34 (9.3) |
| Carbamazepine | 136 (6.6) | Oxcarbazepine | 14 (3.8) |
| Oxcarbazepine | 85 (4.1) | Perampanel | 10 (2.7) |
| Phenytoin | 59 (2.9) | Lacosamide | 9 (2.5) |
| Lacosamide | 42 (2) | Cannabidiol | 9 (2.5) |
| Zonisamide | 24 (1.2) | Zonisamide | 7 (1.9) |
| Perampanel | 22 (1.1) | Primidone | 7 (1.9) |
| - | - | Phenytoin | <5 (0.2) |
| - | - | Ethosuximide | <5 (0.2) |
| ***Antihistamines n (%)*** | ***1445 (1)*** | ***Antihistamines*** | ***191 (1.1)*** |
| Cetirizine | 1111 (76.9) | Cetirizine | 94 (49.2) |
| Diphenhydramine | 121 (8.4) | Levocetirizine | 34 (17.8) |
| Fexofenadine | 56 (3.9) | Promethazine | 13 (6.8) |
| Levocetirizine | 55 (3.8) | Diphenhydramine | 11 (5.8) |
| Promethazine | 25 (1.7) | Doxylamine | 11 (5.8) |
| loratadine | 24 (1.7) | Dimenhydrinate | 7 (3.7) |
| **-** | **-** | Fexofenadine | 5 (2.6) |
| **-** | **-** | Desloratadine | 5 (2.6) |
| **-** | **-** | loratadine | <5 (2.1) |
| **-** | **-** | Cyclizine | <5 (2.1) |
| ***Psychostimulants n (%)*** | ***1389 (0.9)*** | ***Psychostimulants*** | ***192 (1.1)*** |
| Methylphenidate | 469 (33.8) | Methylphenidate | 91 (47.4) |
| Lisdexamfetamine | 279 (20.1) | Methylenedioxymethamphetamine | 30 (15.6) |
| Amphetamine dextroamphetamine | 269 (19.4) | Lisdexamfetamine | 18 (9.4) |
| Atomoxetine | 150 (10.8) | Atomoxetine | 13 (6.8) |
| Modafinil | 55 (4) | Dexamfetamine | 11 (5.7) |
| Armodafinil | 48 (3.5) | Amfetamine | 10 (5.2) |
| Caffeine | 39 (2.8) | Caffeine | 7 (3.6) |
| Dextroamphetamine | 36 (2.6) | Modafinil | 6 (3.1) |
| Dexmethylphenidate | 31 (2.2) | Metamfetamine | <5 (2.0) |
| ***Corticosteroids n (%)*** | ***1167 (0.8)*** | ***Corticosteroids*** | ***317 (1.8)*** |
| Prednisone | 250 (21.4) | Prednisolone | 72 (22.7) |
| Prednisolone | 183 (15.7) | Betamethasone | 62 (19.6) |
| Hydrocortisone | 177 (15.2) | Hydrocortisone | 52 (16.4) |
| Betamethasone | 133 (11.4) | Mometasone | 46 (14.5) |
| Mometasone | 130 (11.1) | Prednisone | 26 (8.2) |
| Triamcinolone | 105 (9) | Dexamethasone | 19 (6) |
| Dexamethasone | 98 (8.4) | Triamcinolone | 14 (4.4) |
| Fluticasone | 49 (4.2) | Fluticasone | 14 (4.4) |
| Budesonide | 33 (2.8) | Budesonide | 8 (2.5) |
| ***Antihypertensives n (%)*** | ***992 (0.7)*** | ***Antihypertensives*** | ***193 (1.1)*** |
| Clonidine | 323 (32.6) | Clonidine | 31 (16.1) |
| Metoprolol | 79 (8) | Metoprolol | 21 (10.9) |
| Propranolol | 79 (8) | Guanfacine | 17 (8.8) |
| Amlodipine | 56 (5.6) | Propranolol | 16 (8.3) |
| Atenolol | 53 (5.3) | bisoprolol | 14 (7.3) |
| Guanfacine | 42 (4.2) | Amlodipine | 10 (5.2) |
| Lisinopril | 39 (3.9) | Ramipril | 8 (4.1) |
| Furosemide | 38 (3.8) | Candesartan | 7 (3.6) |
| bisoprolol | 23 (2.3) | Atenolol | 5 (2.6) |
| Carvedilol | 22 (2.2) | Lisinopril | 5 (2.6) |
| Hydrochlorothiazide | 20 (2) | Furosemide | <5 (2.1) |
| Prazosin | 19 (1.9) | Sotalol | <5 (2.1) |
| Valsartan | 19 (1.9) | Tamsulosin | <5 (2.1) |
| Verapamil | 16 (1.6) | Verapamil | <5 (2.1) |
| Spironolactone | 13 (1.3) | Acetazolamide | <5 (2.1) |
| Nifedipine | 12 (1.2) | Doxazosin | <5 (2.1) |
| Doxazosin | 11 (1.1) | Valsartan | <5 (2.1) |
| Candesartan | 11 (1.1) | Nifedipine | <5 (2.1) |
| Tamsulosin | 10 (1) | Diltiazem | <5 (2.1) |
| Losartan | 10 (1) | - | - |
| ***Immunosuppressants n (%)*** | ***836 (0.6)*** | ***Immunosuppressants*** | ***54 (0.3)*** |
| Natalizumab | 164 (19.6) | Etanercept | 14 (25.9) |
| Tofacitinib | 123 (14.7) | Fingolimod | 11 (20.4) |
| Adalimumab | 122 (14.6) | Adalimumab | 10 (18.5) |
| Infliximab | 72 (8.6) | Dimethyl fumarate | 5 (9.3) |
| Fingolimod | 68 (8.1) | Tofacitinib | <5 (7.4) |
| Etanercept | 54 (6.5) | Infliximab | <5 (7.4) |
| Secukinumab | 51 (6.1) | Secukinumab | <5 (7.4) |
| Dimethyl fumarate | 45 (5.4) | Lenalidomide | <5 (7.4) |
| Lenalidomide | 29 (3.5) | Glatiramer | <5 (7.4) |
| Teriflunomide | 29 (3.5) | - | - |
| Ocrelizumab | 23 (2.8) | - | - |
| Canakinumab | 20 (2.4) | - | - |
| Glatiramer | 19 (2.3) | - | - |
| Abatacept | 17 (2) | - | - |
| ***Anaesthetics n (%)*** | ***809 (0.5)*** | ***Anaesthetics*** | ***290 (1.6)*** |
| Sodium oxybate | 246 (30.4) | Cocaine | 157 (54.1) |
| Cocaine | 186 (23) | Sufentanil | 42 (14.5) |
| Bupivacaine | 153 (18.9) | Ketamine | 40 (13.8) |
| Propofol | 64 (7.9) | Propofol | 18 (6.2) |
| Ketamine | 54 (6.7) | Remifentanil | 9 (3.1) |
| Esketamine | 34 (4.2) | Sodium oxybate | 8 (2.8) |
| Sufentanil | 29 (3.6) | Nitrous oxide | 6 (2.1) |
| Lidocaine | 24 (3) | Capsaicin | <5 (1.3) |
| ***Other analgesics and antipyretics n (%)*** | ***555 (0.4)*** | ***Other analgesics and antipyretics*** | ***95 (0.5)*** |
| Cannabinoids | 194 (35) | Acetaminophen | 56 (58.9) |
| Acetaminophen | 172 (31) | Acetylsalicylic acid | 14 (14.7) |
| Acetaminophen, acetylsalicylic acid and caffeine | 87 (15.7) | Nefopam | 14 (14.7) |
| Ziconotide | 53 (9.5) | Metamizole | 7 (7.4) |
| Acetylsalicylic acid | 38 (6.8) | Ziconotide | <5 (4.2) |
| ***Hormones and related agents n (%)*** | ***504 (0.3)*** | ***Hormones and related agents*** | ***149 (0.8)*** |
| Estradiol | 89 (17.7) | Estradiol | 79 (53) |
| Estrogens medroxyprogesterone | 80 (15.9) | Levonorgestrel | 40 (26.8) |
| Somatropin | 74 (14.7) | Levothyroxine | 10 (6.7) |
| Levothyroxine | 52 (10.3) | Drospirenone estradiol | 5 (3.4) |
| Octreotide | 41 (8.1) | Ethinylestradiol | <5 (2.7) |
| Teriparatide | 39 (7.7) | Somatropin | <5 (2.7) |
| Drospirenone estradiol | 39 (7.7) | Estrogens medroxyprogesterone | <5 (2.7) |
| Levonorgestrel | 37 (7.3) | Medroxyprogesterone | <5 (2.7) |
| Medroxyprogesterone | 20 (4) | - | - |
| Corticotropin | 20 (4) | - | - |
| Megestrol | 13 (2.6) |  |  |
| ***Anti-parkinson drugs n (%)*** | ***504 (0.3)*** | ***Anti-parkinson drugs*** | ***194 (1.1)*** |
| Pramipexole | 157 (31.2) | Pramipexole | 57 (29.4) |
| Levodopa decarboxylase inhibitor | 87 (17.3) | Rasagiline | 35 (18) |
| Amantadine | 63 (12.5) | Levodopa | 24 (12.4) |
| Rotigotine | 47 (9.3) | Levodopa decarboxylase inhibitor | 19 (9.8) |
| Levodopa | 44 (8.7) | Rotigotine | 11 (5.7) |
| Rasagiline | 38 (7.5) | Amantadine | 10 (5.2) |
| Selegiline | 21 (4.2) | Entacapone | 7 (3.6) |
| Tropatepine | 16 (3.2) | Tropatepine | 6 (3.1) |
| Procyclidine | 13 (2.6) | Biperiden | 5 (2.6) |
| Trihexyphenidyl | 5 (1) | Trihexyphenidyl | <5 (2.1) |
| - | - | Carbidopa | <5 (2.1) |
| - | - | Melevodopa | <5 (2.1) |
| - | - | Selegiline | <5 (2.1) |
|  |  | Procyclidine | <5 (2.1) |
| ***Antiinflammatory and antirheumatic products, non-steroids n (%)*** | ***418 (0.28)*** | ***Antiinflammatory and antirheumatic products, non-steroids*** | ***69 (0.4)*** |
| Ibuprofen | 123 (29.4) | Ibuprofen | 37 (53.6) |
| Celecoxib | 97 (23.2) | Diclofenac | 10 (14.5) |
| Rofecoxib | 60 (14.4) | Ketoprofen | 8 (11.6) |
| Naproxen | 43 (10.3) | Etoricoxib | 6 (8.7) |
| Diclofenac | 36 (8.6) | Naproxen | <5 (5.8) |
| Meloxicam | 14 (3.3) | Mefenamic acid | <5 (5.8) |
| Ketorolac | 12 (2.9) | Celecoxib | <5 (5.8) |
| Ketoprofen | 9 (2.2) | **-** | **-** |
| Piroxicam | 8 (1.9) | **-** | **-** |
| Valdecoxib | 5 (1.2) | **-** | **-** |
| Nabumetone | 5 (1.2) | **-** | **-** |
| Mefenamic acid | <5 (1.0) | **-** | **-** |
| ***Antineoplastic agents n (%)*** | ***374 (0.25)*** | ***Antineoplastic agents*** | ***70 (0.4)*** |
| Ruxolitinib | 132 (35.3) | Ruxolitinib | 18 (25.7) |
| Methotrexate | 48 (12.8) | Methotrexate | 13 (18.6) |
| Nilotinib | 32 (8.6) | Imatinib | 11 (15.7) |
| Ibrutinib | 31 (8.3) | Nilotinib | 9 (12.9) |
| Imatinib | 30 (8) | Ibrutinib | <5 (5.7) |
| Palbociclib | 27 (7.2) | Cyclophosphamide | <5 (5.7) |
| Capecitabine | 17 (4.5) | Melphalan | <5 (5.7) |
| doxorubicin | 11 (2.9) | Fluorouracil | <5 (5.7) |
| Dasatinib | 10 (2.7) | Palbociclib | <5 (5.7) |
| Melphalan | 9 (2.4) | Capecitabine | <5 (5.7) |
| Cyclophosphamide | 8 (2.1) | Dasatinib | <5 (5.7) |
| Paclitaxel | 7 (1.9) | Paclitaxel | <5 (5.7) |
| Vincristine | <5 (1.1) | Vincristine | <5 (5.7) |
| Fluorouracil | <5 (1.1) | Etoposide | <5 (5.7) |
| ***Proton pump inhibitors n (%)*** | ***295 (0.2)*** | ***Proton pump inhibitors*** | ***48 (0.27)*** |
| Omeprazole | 129 (43.7) | Omeprazole | 25 (52.1) |
| Esomeprazole | 67 (22.7) | Pantoprazole | 11 (22.9) |
| Pantoprazole | 50 (16.9) | Lansoprazole | 6 (12.5) |
| Lansoprazole | 30 (10.2) | Esomeprazole | <5 (8.3) |
| Rabeprazole | 11 (3.7) | Rabeprazole | <5 (8.3) |
| Dexlansoprazole | 8 (2.7) | - | - |
| ***Antivirals n (%)*** | ***260 (0.17)*** | ***Antivirals*** | ***38 (0.2)*** |
| Ribavirin | 75 (28.8) | Ritonavir | 5 (13.2) |
| Emtricitabine tenofovir | 35 (13.5) | Ribavirin | <5 (10.5) |
| Sofosbuvir | 22 (8.5) | Emtricitabine tenofovir | <5 (10.5) |
| Ledipasvir sofosbuvir | 19 (7.3) | Nevirapine | <5 (10.5) |
| Telaprevir | 17 (6.5) | Nevirapine | <5 (10.5) |
| Atazanavir | 13 (5) | Telaprevir | <5 (10.5) |
| Zidovudine | 10 (3.8) | Ledipasvir sofosbuvir | <5 (10.5) |
| Oseltamivir | 6 (2.3) | Atazanavir | <5 (10.5) |
| Emtricitabine | 6 (2.3) | Simeprevir | <5 (10.5) |
| Oseltamivir | 6 (2.3) | Boceprevir | <5 (10.5) |
| Nevirapine | 5 (1.9) | Aciclovir | <5 (10.5) |
| Simeprevir | 5 (1.9) | Lamivudine | <5 (10.5) |
| Ritonavir | 5 (1.9) | Abacavir | <5 (10.5) |
| Nevirapine | 5 (1.9) | - | - |
| Boceprevir | <5 (1.5) | - | - |
| ***Drugs for constipation n (%)*** | 239 (0.16) | ***Drugs for constipation*** | 34 (0.2) |
| Naloxone | 166 (69.5) | Naloxone | 24 (70.6) |
| Methylnaltrexone | 29 (12.1) | Naldemedine | <5 (11.8) |
| Naldemedine | 28 (11.7) | Bisacodyl | <5 (11.8) |
| Docusate | 9 (3.8) | Senna glycosides | <5 (11.8) |
| Bisacodyl | <5 (1.7) | Methylnaltrexone | <5 (11.8) |
| Senna glycosides | <5 (1.7) | - | - |
| Alvimopan | <5 (1.7) | - | - |
| ***Antibacterials n (%)*** | 237 (0.16) | ***Antibacterials*** | 66 (0.37) |
| Ciprofloxacin | 32 (13.5) | Ciprofloxacin | 7 (10.6) |
| Linezolid | 18 (7.6) | Trimethoprim | 6 (9.1) |
| Azithromycin | 15 (6.3) | Amoxicillin and clavulanic acid | 5 (7.6) |
| Metronidazole | 14 (5.9) | Clarithromycin | 5 (7.6) |
| Amoxicillin | 13 (5.5) | Fusidic acid | 5 (7.6) |
| Trimethoprim | 13 (5.5) | Sulfamethoxazole and trimethoprim (Co-trimoxazole) | <5 (6.1) |
| Vancomycin | 13 (5.5) | Metronidazole | <5 (6.1) |
| Doxycycline | 12 (5.1) | Piperacillin and tazobactam | <5 (6.1) |
| Sulfamethoxazole and trimethoprim (Co-trimoxazole) | 12 (5.1) | Cefuroxime | <5 (6.1) |
| Amoxicillin and clavulanic acid | 11 (4.6) | Doxycycline | <5 (6.1) |
| Clarithromycin | 10 (4.2) | Ofloxacin | <5 (6.1) |
| Piperacillin and tazobactam | 9 (3.8) | Linezolid | <5 (6.1) |
| Cloxacillin | 7 (3) | Vancomycin | <5 (6.1) |
| Moxifloxacin | 7 (3) | Nitrofurantoin | <5 (6.1) |
| Ampicillin and sulbactam | 6 (2.5) | Teicoplanin | <5 (6.1) |
| Nitrofurantoin | 6 (2.5) | Amoxicillin | <5 (6.1) |
| Meropenem | 5 (2.1) | Cloxacillin | <5 (6.1) |
| Levofloxacin | 5 (2.1) | Piperacillin | <5 (6.1) |
| Ceftriaxone | <5 (1.7) | Ceftriaxone | <5 (6.1) |
| Cefpodoxime | <5 (1.7) | Cefixime | <5 (6.1) |
| Aztreonam | <5 (1.7) | Ceftazidime | <5 (6.1) |
| Ofloxacin | <5 (1.7) | Azithromycin | <5 (6.1) |
| ***Urologicals (n = 220)*** | 220 (0.15) | ***Urologicals (n = 36)*** | 36 (0.2) |
| Finasteride | 130 (59.1) | Finasteride | 25 (69.4) |
| Sildenafil | 40 (18.2) | Oxybutynin | <5 (11.1) |
| Oxybutynin | 30 (13.6) | Dutasteride | <5 (11.1) |
| Tadalafil | 14 (6.4) | Sildenafil | <5 (11.1) |
| Dutasteride | <5 (1.8) | Tadalafil | <5 (11.1) |
| Solifenacin | <5 (1.8) | Solifenacin | <5 (11.1) |
| Vardenafil | <5 (1.8) | - | - |
| ***Lipid modifying agents n (%)*** | ***159 (0.1)*** | ***Lipid modifying agents*** | ***19 (0.1)*** |
| Atorvastatin | 63 (39.6) | Simvastatin | 12 (63.2) |
| Simvastatin | 46 (28.9) | Atorvastatin | <5 (21.1) |
| Rosuvastatin | 41 (25.8) | Pravastatin | <5 (21.1) |
| Pravastatin | 9 (5.7%) | - | - |
| ***Anti-dementia drugs n (%)*** | ***146 (0.1)*** | ***Anti-dementia drugs*** | ***46 (0.26)*** |
| Donepezil | 65 (44.5) | Donepezil | 19 (41.3) |
| Memantine | 37 (25.3) | Memantine | 16 (34.8) |
| Galantamine | 23 (15.8) | Rivastigmine | 6 (13) |
| Rivastigmine | 21 (14.4) | Galantamine | 5 (10.9) |
| ***Blood glucose lowering drugs n (%)*** | ***130 (0.08)*** | ***Blood glucose lowering drugs*** | ***29 (0.16)*** |
| Metformin | 37 (28.5) | Metformin | 7 (24.1) |
| Insulin (human) | 20 (15.4) | Insulin (human) | 5 (17.2) |
| Liraglutide | 11 (8.5) | Insulin glargine | 5 (17.2) |
| Sitagliptin | 10 (7.7) | Pioglitazone | <5 (13.8) |
| Insulin glargine | 9 (6.9) | Dulaglutide | <5 (13.8) |
| Pioglitazone | 8 (6.2) | Semaglutide | <5 (13.8) |
| gliclazide | 6 (4.6) | Insulin glulisine | <5 (13.8) |
| Exenatide | 5 (3.8) | Rosiglitazone | <5 (13.8) |
| Rosiglitazone | <5 (3.1) | Sitagliptin | <5 (13.8) |
| Linagliptin | <5 (3.1) | Liraglutide | <5 (13.8) |
| Dulaglutide | <5 (3.1) | Exenatide | <5 (13.8) |
| Canagliflozin | <5 (3.1) | Dapagliflozin | <5 (13.8) |
| glipizide | <5 (3.1) | - | - |
| Dapagliflozin | <5 (3.1) | - | - |
| Empagliflozin | <5 (3.1) | - | - |
| ***Antimigraine preparations n (%)*** | ***91 (0.06)*** | ***Antimigraine preparations*** | ***51 (0.3)*** |
| Sumatriptan | 19 (20.9) | Oxetorone | 20 (39.2) |
| Galcanezumab | 19 (20.9) | Eletriptan | 8 (15.7) |
| Erenumab | 17 (18.7) | Zolmitriptan | 7 (13.7) |
| Eletriptan | 15 (16.5) | Pizotifen | 6 (11.8) |
| Zolmitriptan | 6 (6.6) | Sumatriptan | 5 (9.8) |
| Dihydroergotamine | 5 (5.5) | Dihydroergotamine | <5 (7.8) |
| Naratriptan | <5 (4.4) | Naratriptan | <5 (7.8) |
| Pizotifen | <5 (4.4) | Galcanezumab | <5 (7.8) |
| Methysergide | <5 (4.4) | - | - |
| Rizatriptan | <5 (4.4) | - | - |
| Oxetorone | <5 (4.4) | - | - |
| ***Antimycotics n (%)*** | ***44 (0.03)*** | ***Antimycotics*** | ***7 (0.03)*** |
| Fluconazole | 18 (40.9) | Ketoconazole | <5 (57.1%) |
| Voriconazole | 9 (20.5) | Fluconazole | <5 (57.1%) |
| Ketoconazole | <5 (9.1) | Caspofungin | <5 (57.1%) |
| Caspofungin | <5 (9.1) | itraconazole | <5 (57.1%) |
| itraconazole | <5 (9.1) | Amphotericin B | <5 (57.1%) |
| Posaconazole | <5 (9.1) | Miconazole | <5 (57.1%) |
| Amphotericin B | <5 (9.1) | - | - |
| Miconazole | <5 (9.1) | - | - |

**Table S5.2:** Top 200 indications in FDA-FAERS database

| **No.** | **Indication** | **Frequency** | **Percentage (including unknown indication)**  (*n* = 112,273) | **Percentage (excluding unknown indication)**  (*n* = 55,571) |
| --- | --- | --- | --- | --- |
| 1. | Unknown indication | 56702 | 50.5 | - |
| 2. | Pain | 10399 | 9.26 | 18.72 |
| 3. | Depression | 9080 | 8.09 | 16.34 |
| 4. | Anxiety | 5018 | 4.47 | 9.03 |
| 5. | Substance Use Disorder | 3428 | 3.05 | 6.17 |
| 6. | Muscle Spasticity | 2526 | 2.25 | 4.55 |
| 7. | Neuropathy | 2099 | 1.87 | 3.78 |
| 8. | Fibromyalgia | 1677 | 1.49 | 3.02 |
| 9. | Epilepsy | 1134 | 1.01 | 2.04 |
| 10. | Insomnia | 1015 | 0.9 | 1.83 |
| 11. | Bipolar Disorder | 962 | 0.86 | 1.73 |
| 12. | Schizophrenia | 688 | 0.61 | 1.24 |
| 13. | Smoking Cessation Therapy | 673 | 0.6 | 1.21 |
| 14. | Eczema | 628 | 0.56 | 1.13 |
| 15. | Attention Deficit/Hyperactivity Disorder | 537 | 0.48 | 0.97 |
| 16. | Migraine | 504 | 0.45 | 0.91 |
| 17. | Multiple Sclerosis | 497 | 0.44 | 0.89 |
| 18. | Allergy | 416 | 0.37 | 0.75 |
| 19. | Arthralgia | 334 | 0.3 | 0.60 |
| 20. | Parkinson's Disease | 307 | 0.27 | 0.55 |
| 21. | Post-Traumatic Stress Disorder | 305 | 0.27 | 0.55 |
| 22. | Hypertension | 305 | 0.27 | 0.55 |
| 23. | Cerebral Palsy | 277 | 0.25 | 0.50 |
| 24. | Obsessive-Compulsive Disorder | 227 | 0.2 | 0.41 |
| 25. | Constipation | 219 | 0.2 | 0.39 |
| 26. | Multiple Allergies | 217 | 0.19 | 0.39 |
| 27. | Rheumatoid Arthritis | 212 | 0.19 | 0.38 |
| 28. | Neck Pain | 202 | 0.18 | 0.36 |
| 29. | Headache | 200 | 0.18 | 0.36 |
| 30. | Arthritis | 189 | 0.17 | 0.34 |
| 31. | Nerve Injury | 181 | 0.16 | 0.33 |
| 32. | Analgesic Therapy | 171 | 0.15 | 0.31 |
| 33. | Asthma | 169 | 0.15 | 0.30 |
| 34. | Restless Legs Syndrome | 156 | 0.14 | 0.28 |
| 35. | Osteoarthritis | 145 | 0.13 | 0.26 |
| 36. | Cancer | 145 | 0.13 | 0.26 |
| 37. | Diabetes | 133 | 0.12 | 0.24 |
| 38. | Mental Disorder | 127 | 0.11 | 0.23 |
| 39. | Alcoholism | 122 | 0.11 | 0.22 |
| 40. | Stress | 117 | 0.1 | 0.21 |
| 41. | Intervertebral Disc Degeneration | 115 | 0.1 | 0.21 |
| 42. | Back Injury | 111 | 0.1 | 0.20 |
| 43. | Hepatitis C | 104 | 0.09 | 0.19 |
| 44. | Sedation | 103 | 0.09 | 0.19 |
| 45. | Sciatica | 103 | 0.09 | 0.19 |
| 46. | Intervertebral Disc Protrusion | 97 | 0.09 | 0.17 |
| 47. | Affective Disorder | 97 | 0.09 | 0.17 |
| 48. | Overdose | 94 | 0.08 | 0.17 |
| 49. | Hot Flush | 93 | 0.08 | 0.17 |
| 50. | Prophylaxis | 88 | 0.08 | 0.16 |
| 51. | Osteoporosis | 88 | 0.08 | 0.16 |
| 52. | Social Phobia | 84 | 0.07 | 0.15 |
| 53. | Post Laminectomy Syndrome | 78 | 0.07 | 0.14 |
| 54. | Gastrooesophageal Reflux Disease | 75 | 0.07 | 0.13 |
| 55. | HIV Infection | 75 | 0.07 | 0.13 |
| 56. | Myalgia | 73 | 0.07 | 0.13 |
| 57. | Sedative Therapy | 73 | 0.07 | 0.13 |
| 58. | Fatigue | 70 | 0.06 | 0.13 |
| 59. | Off Label Use | 69 | 0.06 | 0.12 |
| 60. | Crohn's Disease | 67 | 0.06 | 0.12 |
| 61. | Injury | 66 | 0.06 | 0.12 |
| 62. | Agitation | 65 | 0.06 | 0.12 |
| 63. | Contraception | 65 | 0.06 | 0.12 |
| 64. | Abdominal Pain | 64 | 0.06 | 0.12 |
| 65. | Psoriasis | 62 | 0.06 | 0.11 |
| 66. | Alopecia | 59 | 0.05 | 0.11 |
| 67. | Mood Swings | 59 | 0.05 | 0.11 |
| 68. | Surgery | 56 | 0.05 | 0.10 |
| 69. | Irritable Bowel Syndrome | 56 | 0.05 | 0.10 |
| 70. | Menopausal Symptoms | 55 | 0.05 | 0.10 |
| 71. | Narcolepsy | 55 | 0.05 | 0.10 |
| 72. | Rhinitis Allergic | 54 | 0.05 | 0.10 |
| 73. | Systemic Lupus Erythematosus | 54 | 0.05 | 0.10 |
| 74. | Dementia Alzheimer's Type | 54 | 0.05 | 0.10 |
| 75. | Foetal Exposure During Pregnancy | 53 | 0.05 | 0.10 |
| 76. | Nausea | 52 | 0.05 | 0.09 |
| 77. | Premenstrual Syndrome | 50 | 0.04 | 0.09 |
| 78. | Hormone Replacement Therapy | 50 | 0.04 | 0.09 |
| 79. | Psoriatic Arthropathy | 49 | 0.04 | 0.09 |
| 80. | Back Disorder | 48 | 0.04 | 0.09 |
| 81. | Mania | 48 | 0.04 | 0.09 |
| 82. | Herpes Zoster | 48 | 0.04 | 0.09 |
| 83. | Menopause | 48 | 0.04 | 0.09 |
| 84. | Chronic Myeloid Leukaemia | 48 | 0.04 | 0.09 |
| 85. | Lumbar Radiculopathy | 48 | 0.04 | 0.09 |
| 86. | Drug Exposure During Pregnancy | 47 | 0.04 | 0.08 |
| 87. | Borderline Personality Disorder | 47 | 0.04 | 0.08 |
| 88. | Hallucination | 44 | 0.04 | 0.08 |
| 89. | Brain Injury | 43 | 0.04 | 0.08 |
| 90. | Motion Sickness | 42 | 0.04 | 0.08 |
| 91. | Colitis Ulcerative | 41 | 0.04 | 0.07 |
| 92. | Rash | 41 | 0.04 | 0.07 |
| 93. | Drug Therapy | 40 | 0.04 | 0.07 |
| 94. | Urticaria | 40 | 0.04 | 0.07 |
| 95. | Dyspepsia | 39 | 0.03 | 0.07 |
| 96. | Somnolence | 39 | 0.03 | 0.07 |
| 97. | Chronic Hepatitis C | 39 | 0.03 | 0.07 |
| 98. | Ankylosing Spondylitis | 38 | 0.03 | 0.07 |
| 99. | Myelofibrosis | 38 | 0.03 | 0.07 |
| 100. | Agoraphobia | 38 | 0.03 | 0.07 |
| 101. | Spinal Operation | 36 | 0.03 | 0.06 |
| 102. | Paraesthesia | 36 | 0.03 | 0.06 |
| 103. | Cerebrovascular Accident | 35 | 0.03 | 0.06 |
| 104. | Ex-Tobacco User | 35 | 0.03 | 0.06 |
| 105. | Pruritus | 35 | 0.03 | 0.06 |
| 106. | Atrial Fibrillation | 35 | 0.03 | 0.06 |
| 107. | Head Injury | 35 | 0.03 | 0.06 |
| 108. | Chronic Obstructive Pulmonary Disease | 34 | 0.03 | 0.06 |
| 109. | Spinal Osteoarthritis | 33 | 0.03 | 0.06 |
| 110. | Spinal Column Stenosis | 32 | 0.03 | 0.06 |
| 111. | Weight Decreased | 32 | 0.03 | 0.06 |
| 112. | Aggression | 32 | 0.03 | 0.06 |
| 113. | Road Traffic Accident | 31 | 0.03 | 0.06 |
| 114. | Androgenetic Alopecia | 31 | 0.03 | 0.06 |
| 115. | Joint Injury | 30 | 0.03 | 0.05 |
| 116. | Suicidal Ideation | 30 | 0.03 | 0.05 |
| 117. | Panic Reaction | 29 | 0.03 | 0.05 |
| 118. | Radiculopathy | 29 | 0.03 | 0.05 |
| 119. | Diarrhoea | 28 | 0.02 | 0.05 |
| 120. | Tobacco User | 28 | 0.02 | 0.05 |
| 121. | Bone Pain | 27 | 0.02 | 0.05 |
| 122. | Dizziness | 27 | 0.02 | 0.05 |
| 123. | Metastases To Bone | 27 | 0.02 | 0.05 |
| 124. | Delirium | 27 | 0.02 | 0.05 |
| 125. | Hypothyroidism | 27 | 0.02 | 0.05 |
| 126. | Cataplexy | 26 | 0.02 | 0.05 |
| 127. | Hypercholesterolaemia | 26 | 0.02 | 0.05 |
| 128. | Nervous System Disorder | 26 | 0.02 | 0.05 |
| 129. | Euphoric Mood | 25 | 0.02 | 0.04 |
| 130. | Adverse Drug Reaction | 25 | 0.02 | 0.04 |
| 131. | Urinary Tract Infection | 25 | 0.02 | 0.04 |
| 132. | Postoperative Analgesia | 25 | 0.02 | 0.04 |
| 133. | Dystonia | 24 | 0.02 | 0.04 |
| 134. | Intentional Product Misuse | 24 | 0.02 | 0.04 |
| 135. | Chronic Fatigue Syndrome | 24 | 0.02 | 0.04 |
| 136. | Plasma Cell Myeloma | 24 | 0.02 | 0.04 |
| 137. | Nerve Compression | 23 | 0.02 | 0.04 |
| 138. | Mood Altered | 23 | 0.02 | 0.04 |
| 139. | Endometriosis | 22 | 0.02 | 0.04 |
| 140. | Maternal Exposure Timing Unspecified | 22 | 0.02 | 0.04 |
| 141. | Scoliosis | 22 | 0.02 | 0.04 |
| 142. | Alcohol Use | 22 | 0.02 | 0.04 |
| 143. | Vomiting | 22 | 0.02 | 0.04 |
| 144. | Intervertebral Disc Disorder | 22 | 0.02 | 0.04 |
| 145. | Acne | 21 | 0.02 | 0.04 |
| 146. | Tremor | 21 | 0.02 | 0.04 |
| 147. | Nervousness | 21 | 0.02 | 0.04 |
| 148. | Blood Cholesterol Increased | 21 | 0.02 | 0.04 |
| 149. | Tardive Dyskinesia | 21 | 0.02 | 0.04 |
| 150. | Infection | 21 | 0.02 | 0.04 |
| 151. | Catatonia | 21 | 0.02 | 0.04 |
| 152. | Abdominal Pain Upper | 21 | 0.02 | 0.04 |
| 153. | Suicide Attempt | 20 | 0.02 | 0.04 |
| 154. | Psychomotor Hyperactivity | 20 | 0.02 | 0.04 |
| 155. | General Anaesthesia | 20 | 0.02 | 0.04 |
| 156. | Dementia | 20 | 0.02 | 0.04 |
| 157. | Limb Injury | 20 | 0.02 | 0.04 |
| 158. | Quadriplegia | 20 | 0.02 | 0.04 |
| 159. | Anger | 19 | 0.02 | 0.03 |
| 160. | Gout | 19 | 0.02 | 0.03 |
| 161. | Chronic Lymphocytic Leukaemia | 19 | 0.02 | 0.03 |
| 162. | Tendonitis | 19 | 0.02 | 0.03 |
| 163. | Muscle Relaxant Therapy | 18 | 0.02 | 0.03 |
| 164. | Myofascial Pain Syndrome | 18 | 0.02 | 0.03 |
| 165. | Pelvic Pain | 18 | 0.02 | 0.03 |
| 166. | Pneumonia | 18 | 0.02 | 0.03 |
| 167. | Weight Control | 18 | 0.02 | 0.03 |
| 168. | Autism Spectrum Disorder | 18 | 0.02 | 0.03 |
| 169. | Nasal Congestion | 18 | 0.02 | 0.03 |
| 170. | Drug Detoxification | 17 | 0.02 | 0.03 |
| 171. | Asthenia | 17 | 0.02 | 0.03 |
| 172. | Knee Arthroplasty | 17 | 0.02 | 0.03 |
| 173. | Stress Symptoms | 17 | 0.02 | 0.03 |
| 174. | Radicular Pain | 17 | 0.02 | 0.03 |
| 175. | Gait Disturbance | 17 | 0.02 | 0.03 |
| 176. | Spinal Disorder | 16 | 0.01 | 0.03 |
| 177. | Nerve Block | 16 | 0.01 | 0.03 |
| 178. | Dysmenorrhoea | 16 | 0.01 | 0.03 |
| 179. | Polymyalgia Rheumatica | 16 | 0.01 | 0.03 |
| 180. | Spasticity | 16 | 0.01 | 0.03 |
| 181. | Decreased Appetite | 16 | 0.01 | 0.03 |
| 182. | Multiple Myeloma | 16 | 0.01 | 0.03 |
| 183. | Chest Pain | 16 | 0.01 | 0.03 |
| 184. | Sinus Disorder | 16 | 0.01 | 0.03 |
| 185. | Spinal Fracture | 15 | 0.01 | 0.03 |
| 186. | Spinal Cord Disorder | 15 | 0.01 | 0.03 |
| 187. | Blood Testosterone Decreased | 15 | 0.01 | 0.03 |
| 188. | Neck Injury | 15 | 0.01 | 0.03 |
| 189. | Spinal Fusion Surgery | 15 | 0.01 | 0.03 |
| 190. | Gastritis | 15 | 0.01 | 0.03 |
| 191. | Sinusitis | 15 | 0.01 | 0.03 |
| 192. | Detoxification | 14 | 0.01 | 0.03 |
| 193. | Cardiac Failure | 14 | 0.01 | 0.03 |
| 194. | Personality Disorder | 14 | 0.01 | 0.03 |
| 195. | Hereditary Ataxia | 14 | 0.01 | 0.03 |
| 196. | Phantom Pain | 14 | 0.01 | 0.03 |
| 197. | Blood Pressure Abnormal | 14 | 0.01 | 0.03 |
| 198. | Erectile Dysfunction | 14 | 0.01 | 0.03 |
| 199. | Dyskinesia | 14 | 0.01 | 0.03 |
| 200. | Neoplasm Malignant | 14 | 0.01 | 0.03 |

**Table S5.3:** Top 200 indications in EMA-EV database

| **No.** | **Indications** | **Frequency** | **Percentage (including unknown indication)**  (*n*= 21,633) | **Percentage (excluding unknown indication)**  (*n* = 18,111) |
| --- | --- | --- | --- | --- |
| 1. | Product Used for Unknown Indication | 3522 | 16.28 | - |
| 2. | Depression | 2151 | 9.94 | 11.87 |
| 3. | Pain | 1474 | 6.81 | 8.13 |
| 4. | Anxiety | 1080 | 4.99 | 5.96 |
| 5. | Drug Dependence | 519 | 2.40 | 2.86 |
| 6. | Neuralgia | 516 | 2.39 | 2.85 |
| 7. | Back Pain | 480 | 2.22 | 2.65 |
| 8. | Insomnia | 405 | 1.87 | 2.24 |
| 9. | Sleep Disorder | 347 | 1.60 | 1.91 |
| 10. | Schizophrenia | 325 | 1.50 | 1.79 |
| 11. | Drug Abuse | 270 | 1.25 | 1.49 |
| 12. | Parkinson's Disease | 263 | 1.22 | 1.45 |
| 13. | Bipolar Disorder | 254 | 1.17 | 1.40 |
| 14. | Smoking Cessation Therapy | 232 | 1.07 | 1.28 |
| 15. | Epilepsy | 228 | 1.05 | 1.26 |
| 16. | Drug Withdrawal Maintenance Therapy | 194 | 0.90 | 1.07 |
| 17. | Eczema | 188 | 0.87 | 1.04 |
| 18. | Anxiety Disorder | 187 | 0.86 | 1.03 |
| 19. | Migraine | 170 | 0.79 | 0.94 |
| 20. | Arthralgia | 156 | 0.72 | 0.86 |
| 21. | Psychotic Disorder | 145 | 0.67 | 0.80 |
| 22. | Sedation | 139 | 0.64 | 0.77 |
| 23. | Panic Attack | 137 | 0.63 | 0.76 |
| 24. | Attention Deficit Hyperactivity Disorder | 130 | 0.60 | 0.72 |
| 25. | Analgesic Therapy | 129 | 0.60 | 0.71 |
| 26. | Panic Disorder | 128 | 0.59 | 0.71 |
| 27. | Generalised Anxiety Disorder | 127 | 0.59 | 0.70 |
| 28. | Major Depression | 124 | 0.57 | 0.68 |
| 29. | Product Substitution | 110 | 0.51 | 0.61 |
| 30. | Procedural Pain | 109 | 0.50 | 0.60 |
| 31. | Fibromyalgia | 101 | 0.47 | 0.56 |
| 32. | Withdrawal Syndrome | 100 | 0.46 | 0.55 |
| 33. | Post-Traumatic Stress Disorder | 98 | 0.45 | 0.54 |
| 34. | Headache | 95 | 0.44 | 0.52 |
| 35. | Depressed Mood | 92 | 0.43 | 0.51 |
| 36. | Contraception | 87 | 0.40 | 0.48 |
| 37. | Restless Legs Syndrome | 86 | 0.40 | 0.47 |
| 38. | Hormone Replacement Therapy | 85 | 0.39 | 0.47 |
| 39. | Obsessive-Compulsive Disorder | 81 | 0.37 | 0.45 |
| 40. | Alcoholism | 79 | 0.37 | 0.44 |
| 41. | Hypertension | 79 | 0.37 | 0.44 |
| 42. | Mental Disorder | 78 | 0.36 | 0.43 |
| 43. | Muscle Spasticity | 76 | 0.35 | 0.42 |
| 44. | Intervertebral Disc Protrusion | 75 | 0.35 | 0.41 |
| 45. | Menopause | 67 | 0.31 | 0.37 |
| 46. | Dermatitis Atopic | 65 | 0.30 | 0.36 |
| 47. | Stress | 65 | 0.30 | 0.36 |
| 48. | Sciatica | 63 | 0.29 | 0.35 |
| 49. | Antidepressant Therapy | 61 | 0.28 | 0.34 |
| 50. | Sedative Therapy | 60 | 0.28 | 0.33 |
| 51. | Schizoaffective Disorder | 58 | 0.27 | 0.32 |
| 52. | Neck Pain | 57 | 0.26 | 0.31 |
| 53. | Asthma | 56 | 0.26 | 0.31 |
| 54. | Mixed Anxiety And Depressive Disorder | 56 | 0.26 | 0.31 |
| 55. | Drug Withdrawal Syndrome | 54 | 0.25 | 0.30 |
| 56. | Borderline Personality Disorder | 53 | 0.24 | 0.29 |
| 57. | Constipation | 50 | 0.23 | 0.28 |
| 58. | Seasonal Allergy | 50 | 0.23 | 0.28 |
| 59. | Off Label Use | 47 | 0.22 | 0.26 |
| 60. | Dementia Alzheimer's Type | 47 | 0.22 | 0.26 |
| 61. | Burnout Syndrome | 46 | 0.21 | 0.25 |
| 62. | Hypersensitivity | 45 | 0.21 | 0.25 |
| 63. | Cancer Pain | 44 | 0.20 | 0.24 |
| 64. | Affective Disorder | 43 | 0.20 | 0.24 |
| 65. | Breakthrough Pain | 42 | 0.19 | 0.23 |
| 66. | Restlessness | 42 | 0.19 | 0.23 |
| 67. | Partial Seizures | 42 | 0.19 | 0.23 |
| 68. | Social Anxiety Disorder | 41 | 0.19 | 0.23 |
| 69. | Tobacco User | 41 | 0.19 | 0.23 |
| 70. | Substance Use | 39 | 0.18 | 0.22 |
| 71. | Prophylaxis | 39 | 0.18 | 0.22 |
| 72. | Pain In Extremity | 39 | 0.18 | 0.22 |
| 73. | Spinal Pain | 38 | 0.18 | 0.21 |
| 74. | Fatigue | 38 | 0.18 | 0.21 |
| 75. | Bipolar I Disorder | 38 | 0.18 | 0.21 |
| 76. | Adjustment Disorder with Depressed Mood | 38 | 0.18 | 0.21 |
| 77. | Abdominal Pain | 37 | 0.17 | 0.20 |
| 78. | Alcohol Use | 36 | 0.17 | 0.20 |
| 79. | Osteoarthritis | 36 | 0.17 | 0.20 |
| 80. | Rheumatoid Arthritis | 35 | 0.16 | 0.19 |
| 81. | Seizure | 35 | 0.16 | 0.19 |
| 82. | Nausea | 34 | 0.16 | 0.19 |
| 83. | Somatic Symptom Disorder | 34 | 0.16 | 0.19 |
| 84. | Myalgia | 34 | 0.16 | 0.19 |
| 85. | Trigeminal Neuralgia | 34 | 0.16 | 0.19 |
| 86. | Acne | 33 | 0.15 | 0.18 |
| 87. | Foetal Exposure During Pregnancy | 33 | 0.15 | 0.18 |
| 88. | Perinatal Depression | 33 | 0.15 | 0.18 |
| 89. | Bulimia Nervosa | 32 | 0.15 | 0.18 |
| 90. | Agitation | 32 | 0.15 | 0.18 |
| 91. | Intentional Product Misuse | 31 | 0.14 | 0.17 |
| 92. | HIV Infection | 31 | 0.14 | 0.17 |
| 93. | Multiple Sclerosis | 31 | 0.14 | 0.17 |
| 94. | Alcohol Withdrawal Syndrome | 30 | 0.14 | 0.17 |
| 95. | Toxicity To Various Agents | 30 | 0.14 | 0.17 |
| 96. | Substance Abuse | 30 | 0.14 | 0.17 |
| 97. | Migraine Prophylaxis | 28 | 0.13 | 0.15 |
| 98. | Pain Management | 28 | 0.13 | 0.15 |
| 99. | Metastases To Peritoneum | 28 | 0.13 | 0.15 |
| 100. | Generalised Tonic-Clonic Seizure | 28 | 0.13 | 0.15 |
| 101. | Ill-Defined Disorder | 28 | 0.13 | 0.15 |
| 102. | Autism Spectrum Disorder | 27 | 0.12 | 0.15 |
| 103. | Systemic Lupus Erythematosus | 27 | 0.12 | 0.15 |
| 104. | Drug Use Disorder | 27 | 0.12 | 0.15 |
| 105. | Neuropathy Peripheral | 27 | 0.12 | 0.15 |
| 106. | Spondylitis | 26 | 0.12 | 0.14 |
| 107. | Chronic Myeloid Leukaemia | 26 | 0.12 | 0.14 |
| 108. | Polyneuropathy | 25 | 0.12 | 0.14 |
| 109. | Endometriosis | 24 | 0.11 | 0.13 |
| 110. | Neurosis | 24 | 0.11 | 0.13 |
| 111. | Atrial Fibrillation | 24 | 0.11 | 0.13 |
| 112. | Suicide Attempt | 24 | 0.11 | 0.13 |
| 113. | Abdominal Pain Upper | 24 | 0.11 | 0.13 |
| 114. | Alcohol Abuse | 24 | 0.11 | 0.13 |
| 115. | Muscle Spasms | 24 | 0.11 | 0.13 |
| 116. | Alopecia | 23 | 0.11 | 0.13 |
| 117. | Maternal Exposure Timing Unspecified | 22 | 0.10 | 0.12 |
| 118. | Menopausal Symptoms | 21 | 0.10 | 0.12 |
| 119. | Antiplatelet Therapy | 21 | 0.10 | 0.12 |
| 120. | Colitis Ulcerative | 21 | 0.10 | 0.12 |
| 121. | Overdose | 21 | 0.10 | 0.12 |
| 122. | Psoriasis | 21 | 0.10 | 0.12 |
| 123. | Hot Flush | 21 | 0.10 | 0.12 |
| 124. | Complex Regional Pain Syndrome | 21 | 0.10 | 0.12 |
| 125. | Giant Cell Arteritis | 20 | 0.09 | 0.11 |
| 126. | Arthritis | 20 | 0.09 | 0.11 |
| 127. | Herpes Zoster | 20 | 0.09 | 0.11 |
| 128. | Fear | 20 | 0.09 | 0.11 |
| 129. | Dizziness | 20 | 0.09 | 0.11 |
| 130. | Chronic Fatigue Syndrome | 19 | 0.09 | 0.10 |
| 131. | Gastrointestinal Disorder | 19 | 0.09 | 0.10 |
| 132. | Personality Disorder | 19 | 0.09 | 0.10 |
| 133. | Postoperative Analgesia | 19 | 0.09 | 0.10 |
| 134. | Hepatitis C | 18 | 0.08 | 0.10 |
| 135. | Chest Pain | 18 | 0.08 | 0.10 |
| 136. | General Symptom | 18 | 0.08 | 0.10 |
| 137. | Diabetic Neuropathy | 18 | 0.08 | 0.10 |
| 138. | Stress Urinary Incontinence | 18 | 0.08 | 0.10 |
| 139. | Cluster Headache | 18 | 0.08 | 0.10 |
| 140. | Nervousness | 17 | 0.08 | 0.09 |
| 141. | Paraesthesia | 17 | 0.08 | 0.09 |
| 142. | COVID-19 Immunisation | 17 | 0.08 | 0.09 |
| 143. | Failed Back Surgery Syndrome | 17 | 0.08 | 0.09 |
| 144. | Detoxification | 17 | 0.08 | 0.09 |
| 145. | Tinnitus | 17 | 0.08 | 0.09 |
| 146. | Tetany | 17 | 0.08 | 0.09 |
| 147. | Dependence | 16 | 0.07 | 0.09 |
| 148. | Sinus Tachycardia | 16 | 0.07 | 0.09 |
| 149. | Polymyalgia Rheumatica | 16 | 0.07 | 0.09 |
| 150. | Suicidal Ideation | 16 | 0.07 | 0.09 |
| 151. | Facial Pain | 16 | 0.07 | 0.09 |
| 152. | Injury | 15 | 0.07 | 0.08 |
| 153. | Tension Headache | 15 | 0.07 | 0.08 |
| 154. | Relapsing-Remitting Multiple Sclerosis | 15 | 0.07 | 0.08 |
| 155. | Poor Quality Sleep | 15 | 0.07 | 0.08 |
| 156. | Migraine With Aura | 15 | 0.07 | 0.08 |
| 157. | Nerve Injury | 15 | 0.07 | 0.08 |
| 158. | Premenstrual Syndrome | 15 | 0.07 | 0.08 |
| 159. | Plasma Cell Myeloma | 14 | 0.06 | 0.08 |
| 160. | Chronic Hepatitis C | 14 | 0.06 | 0.08 |
| 161. | Toothache | 14 | 0.06 | 0.08 |
| 162. | Supraventricular Extrasystoles | 14 | 0.06 | 0.08 |
| 163. | Hypercholesterolaemia | 14 | 0.06 | 0.08 |
| 164. | Thinking Abnormal | 14 | 0.06 | 0.08 |
| 165. | Irritable Bowel Syndrome | 14 | 0.06 | 0.08 |
| 166. | Exposure Via Breast Milk | 14 | 0.06 | 0.08 |
| 167. | Parkinsonism | 13 | 0.06 | 0.07 |
| 168. | Type 2 Diabetes Mellitus | 13 | 0.06 | 0.07 |
| 169. | Tension | 13 | 0.06 | 0.07 |
| 170. | Heavy Menstrual Bleeding | 13 | 0.06 | 0.07 |
| 171. | Rhinitis Allergic | 13 | 0.06 | 0.07 |
| 172. | Nervous System Disorder | 13 | 0.06 | 0.07 |
| 173. | Nasal Congestion | 13 | 0.06 | 0.07 |
| 174. | Dementia | 13 | 0.06 | 0.07 |
| 175. | Petit Mal Epilepsy | 12 | 0.06 | 0.07 |
| 176. | Psychotic Symptom | 12 | 0.06 | 0.07 |
| 177. | Pneumonia | 12 | 0.06 | 0.07 |
| 178. | Compulsions | 12 | 0.06 | 0.07 |
| 179. | Bipolar II Disorder | 12 | 0.06 | 0.07 |
| 180. | Cystic Fibrosis | 12 | 0.06 | 0.07 |
| 181. | Urticaria | 12 | 0.06 | 0.07 |
| 182. | Osteoporosis | 12 | 0.06 | 0.07 |
| 183. | Dystonia | 12 | 0.06 | 0.07 |
| 184. | Temporal Lobe Epilepsy | 11 | 0.05 | 0.06 |
| 185. | Spinal Stenosis | 11 | 0.05 | 0.06 |
| 186. | Persistent Depressive Disorder | 11 | 0.05 | 0.06 |
| 187. | Foetal Exposure Timing Unspecified | 11 | 0.05 | 0.06 |
| 188. | Hypothyroidism | 11 | 0.05 | 0.06 |
| 189. | Hernia | 11 | 0.05 | 0.06 |
| 190. | Myelofibrosis | 11 | 0.05 | 0.06 |
| 191. | Ehlers-Danlos Syndrome | 11 | 0.05 | 0.06 |
| 192. | Depressive Symptom | 11 | 0.05 | 0.06 |
| 193. | Endotracheal Intubation | 11 | 0.05 | 0.06 |
| 194. | Analgesic Intervention Supportive Therapy | 11 | 0.05 | 0.06 |
| 195. | Postoperative Care | 11 | 0.05 | 0.06 |
| 196. | Hallucination | 11 | 0.05 | 0.06 |
| 197. | Intervertebral Disc Degeneration | 11 | 0.05 | 0.06 |
| 198. | Self-Medication | 11 | 0.05 | 0.06 |
| 199. | Nerve Compression | 10 | 0.05 | 0.06 |
| 200. | Cough | 10 | 0.05 | 0.06 |
