## Supplementary material for "Reporting patterns of adverse drug withdrawal events using individual case safety reports in United States and European databases": Chi-square analysis of ADWE seriousness by age and sex in FDA-FAERS and EMA-EV (Tables S6.1-S6.2)

**Supplementary file 6: Chi-square analysis of seriousness criteria of ADWEs reports by age and sex group in FDA-FAERS and EMA-EV databases (Table S6.1 and S6.2)**

**Chi-square analysis by age group in FDA-FAERS and EMA-EV databases**

***Details:*** Both seriousness criteria and age group information were available for 55,370 and 2,362 cases in the FDA-FAERS and EMA-EV databases, respectively. Significant associations between age group and seriousness criteria were observed for hospitalisation, disability and other serious/important medical condition in both datasets (p < 0.05) with commonly reported in adults and older adults. However, in the EMA database, life-threatening (LT) and death (DE) outcomes showed no significant differences across age groups (p = 0.846 and p = 0.584, respectively). Congenital anomalies were more commonly reported in paediatric cases (FDA-FAERS = 54.9% and EMA-EV = 90%) with a strongest association in both datasets (FDA-FAERS: χ² = 652.36; EMA-EV: χ² = 126.36; p < 0.0001) ***(Table S6.1)****.*

**Table S6.1:** Chi-square analysis by age group in FDA-FAERS and EMA-EV databases

| **FDA-FAERS (55,370)** | | | | | | | | | **EMA-EV (n=2362)** | | | | | | | | |
| --- | --- | --- | --- | --- | --- | --- | --- | --- | --- | --- | --- | --- | --- | --- | --- | --- | --- |
| **Seriousness criteria** | **Age group** | | | | | | **χ² (df)** | **p-value** | **Seriousness criteria** | **Age group** | | | | | | **χ² (df)** | **p-value** |
|  | **Paediatrics** | | **Adult** | | **Older adults** | |  |  |  | **Paediatrics** | | **Adult** | | **Older adults** | |  |  |
| **Outcome** | **Yes**  **(n, %)** | **No**  **(n, %)** | **Yes**  **(n, %)** | **No**  **(n, %)** | **Yes**  **(n, %)** | **No**  **(n, %)** |  |  | **Outcome** | **Yes**  **(n, %)** | **No**  **(n, %)** | **Yes**  **(n, %)** | **No**  **(n, %)** | **Yes**  **(n, %)** | **No**  **(n, %)** | **χ² (df)** | **p-value** |
| Other serious/important medical condition (Yes = 31142; No = 24228) | 2,124 (6.8%) | 990 (4.1%) | 24,692 (79.3%) | 19,141 (79.0%) | 4,326 (13.9%) | 4,097 (16.9%) | 262.92(2) | **0.000** | (Yes = 1054; No = 1308) | 77  (7.3%) | 64 (4.9%) | 743 (70.5%) | 1,049 (80.2%) | 234 (22.2%) | 195 (14.9%) | 30.03(2) | **0.000** |
| Hospitalisation (Yes = 16405; No = 38965) | 1,206 (7.4%) | 1,908 (4.9%) | 12,401 (75.6%) | 31,432 (80.7%) | 2,798 (17.1%) | 5,625 (14.4%) | 213.33(2) | **0.000** | (Yes = 530; No = 1832) | 61  (11.5%) | 80 (4.4%) | 356 (67.2%) | 1,436 (78.4%) | 113 (21.3%) | 316 (17.3%) | 45.70(2) | **0.000** |
| Disability  (Yes = 4323; No = 51047) | 180 (4.2%) | 2,934 (5.7%) | 3,767 (87.1%) | 40,066 (78.5%) | 376 (8.7%) | 8,047 (15.8%) | 186.27(2) | **0.000** | (Yes = 100; No = 2262) | 1  (1.0%) | 140 (6.2%) | 88 (88.0%) | 1,704 (75.3%) | 11 (11.0%) | 418 (18.5%) | 9.30(2) | **0.010** |
| Life-threatening condition (Yes = 2908; No = 52462) | 208 (7.2%) | 2,906 (5.5%) | 2,451 (84.3%) | 41,382 (78.9%) | 249 (8.6%) | 8,174 (15.6%) | 112.13(2) | **0.000** | (Yes = 88; No = 2274) | 4  (4.5%) | 137 (6.0%) | 68 (77.3%) | 1,724 (75.8%) | 16 (18.2%) | 413 (18.2%) | 0.33(2) | 0.846 |
| Death  (Yes= 1409; No = 53961) | 72 (5.1%) | 3,042 (5.6%) | 1,072 (76.1%) | 42,761 (79.2%) | 265 (18.8%) | 8,158 (15.1%) | 14.70(2) | **0.001** | (Yes= 28; No = 2334) | 1  (3.6%) | 140 (6.0%) | 20 (71.4%) | 1,772 (75.9%) | 7  (25.0%) | 422 (18.1%) | 1.08(2) | 0.584 |
| Congenital anomaly (Yes = 142; No = 55228) | 78 (54.9%) | 3,036 (5.5%) | 56 (39.4%) | 43,777 (79.3%) | 8 (5.6%) | 8,415 (15.2%) | 652.36(2) | **0.000** | (Yes = 10; No = 2352) | 9  (90.0%) | 132 (5.6%) | 1  (10.0%) | 1,791 (94.6%) | 0  (0.0%) | 429 (100.0%) | 126.36(2) | **0.000** |
| Required intervention to prevent permanent impairment/damage*(Yes = 54248; No = 1122) | 97 (8.6%) | 3,017 (5.6%) | 954 (85.0%) | 42,879 (79.0%) | 71 (6.3%) | 8,352 (15.4%) | 82.98(2) | **0.000** | - | - | - | - | - | - | - | - | - |

**: Only available in FDA; χ² =Chi-square statistic; df = degree of freedom*

**Chi-square analysis by sex group in FDA-FAERS and EMA-EV databases**

***Details:*** Analysis of the FDA-FAERS (n=134,105) and EMA-EV (n=8,062) datasets also revealed sex-based differences in ADWE seriousness criteria. Analysis of the FDA-FAERS and EMA-EV datasets also revealed sex-based differences in ADWE seriousness criteria. In FDA-FAERS data, there were males reported significantly higher proportions of death for males (62.5% vs. 37.5%, p < 0.001), disability (56.4% vs. 43.6%, p < 0.001) and congenital anomalies (51.2% vs. 48.8%, p = 0.018) compared to females. Females showed a higher proportion of other serious conditions (50.9% vs. 49.1%, p < 0.001). No significant sex differences were observed for hospitalisation (p = 0.930) or life-threatening condition (p = 0.417). In EMA-EV dataset, males had significantly higher rates of hospitalization (52.4% vs. 47.6%, p < 0.001), life-threatening events (52.7% vs. 47.3%, p < 0.001), and death (65.3% vs. 34.7%, p < 0.001) compared to females. However, females had higher reports of disability (63.4% vs. 36.6%, p = 0.004). Moreover, no significant sex differences were observed for other serious conditions (p = 0.233) or congenital anomalies (p = 0.406) ***(Table S6.2)***. These differences may reflect variations in population characteristics, reporting practices, or regulatory frameworks between the FDA-FAERS and EMA-EV.

**Table S6.2:** Chi-square analysis by sex group in FDA-FAERS and EMA-EV databases

| **FDA-FAERS (*n*=134,105)** | | | | | | | **EMA-EV (*n* = 8,062)** | | | | | | |
| --- | --- | --- | --- | --- | --- | --- | --- | --- | --- | --- | --- | --- | --- |
| **Seriousness criteria** | **Sex group** | | | | **χ² (df)** | **p-value** | **Seriousness criteria** | **Sex group** | | | | **χ² (df)** | **p-value** |
|  | **Female** | | **Male** | |  |  |  | **Female** | | **Male** | |  |  |
| **Outcome** | **Yes**  **(n, %)** | **No**  **(n, %)** | **Yes**  **(n, %)** | **No**  **(n, %)** |  |  | **Outcome** | **Yes**  **(n, %)** | **No**  **(n, %)** | **Yes**  **(n, %)** | **No**  **(n, %)** |  |  |
| Other serious/important medical condition (Yes = 90747; No = 43358) | 46,180 (50.9%) | 27,983 (64.5%) | 44,567 (49.1%) | 15,375 (35.5%) | 2200 (1) | **0.000** | (Yes = 3,678; No = 4384) | 2,135 (58.1%) | 2,487 (56.7%) | 1,543 (41.9%) | 1,897 (43.3%) | 1.42 (1) | 0.233 |
| Hospitalisation (Yes = 22773; No = 111332) | 12,600 (55.3%) | 61,563 (55.3%) | 10,173 (44.7%) | 49,769 (44.7%) | 0.007 (1) | 0.930 | (Yes = 2,310; No = 5752) | 1,099 (47.6%) | 3,523 (61.2%) | 1,211 (52.4%) | 2,229 (38.8%) | 125.95 (1) | **0.000** |
| Disability (Yes = 20000; No = 114105) | 8,716 (43.6%) | 65,447 (57.4%) | 11,284 (56.4%) | 48,658 (42.6%) | 1300 (1) | **0.000** | (Yes = 513; No = 7,549) | 325  (63.4%) | 4,297 (56.9%) | 188 (36.6%) | 3,252 (43.1%) | 8.12 (1) | **0.004** |
| Life-threatening condition (Yes = 3613; No = 130492) | 2,022 (56.0%) | 72,141 (55.3%) | 1,591 (44.0%) | 58,351 (44.7%) | 0.66(1) | 0.417 | (Yes = 300; No = 7,762) | 142  (47.3%) | 4,480 (57.7%) | 158 (52.7%) | 3,282 (42.3%) | 12.73 (1) | **0.000** |
| Death (Yes = 4249; No = 129856) | 1,595 (37.5%) | 72,568 (55.9%) | 2,654 (62.5%) | 57,288 (44.1%) | 560.17 (1) | **0.000** | (Yes = 75;  No = 7.987) | 26  (34.7%) | 4,596 (57.5%) | 49  (65.3%) | 3,391 (42.5%) | 15.90 (1) | **0.000** |
| Congenital anomaly (Yes = 320; No = 133785) | 156 (48.8%) | 74,007 (55.3%) | 164 (51.2%) | 59,778 (44.7%) | 5.57 (1) | **0.018** | (Yes = 26;  No = 8036) | 17  (65.4%) | 4,605 (57.3%) | 9  (34.6%) | 3,431 (42.7%) | 0.69 (1) | 0.406 |
| Required intervention to prevent permanent impairment/damage*(Yes = 1768; No = 132337) | 936 (52.9%) | 73,227 (55.3%) | 832 (47.1%) | 59,110 (44.7%) | 4.04 (1) | **0.044** | - | - | - | - | - | - | - |

**: Only available in FDA-FAERS; χ² =Chi-square statistic; df = degree of freedom*
